# Small or large patches? History, biophysics, and an empirical test of a core assumption in EEG independent component analysis

**DOI:** 10.64898/2026.01.23.26344529

**Authors:** Komal K. Kukkar, Hyeonseok Kim, Pranav J. Parikh, Makoto Miyakoshi

**Affiliations:** Center for Neuromotor and Biomechanics Research, Department of Health and Human Performance, University of Houston, Houston, TX 77004-6015; Division of Child and Adolescent Psychiatry, Cincinnati Children’s Hospital Medical Center, Cincinnati, OH 45229; Department of Psychiatry and Behavioral Neuroscience, University of Cincinnati College of Medicine, Cincinnati, OH 45219-0559

**Keywords:** EEG, independent component analysis (ICA), dipole, source localization, volume conduction

## Abstract

This is a methods paper on the physiological interpretation of independent component analysis (ICA) applied to scalp EEG, but its scope extends to historical review and hypothesis-driven research. First, we trace how the small-patch model, which equates each brain independent component (IC) with a cortical patch smaller than 1 cm², became the central assumption of this interpretation. Second, we review a clinically and biophysically grounded alternative, the large-patch model (≥ 6 cm²). The two models make opposing predictions: the small-patch model predicts that about two-thirds of ICs lie on sulcal walls with tangential dipoles, whereas the large-patch model predicts radial dipoles displaced deeper than the cortex because fitting a single dipole to an extended source produces depth bias. Third, we test them: in simulation, depth bias grew with patch radius and was modulated, but not eliminated, by skull-conductivity mismatch. In resting-state EEG from two open datasets (820 recordings; 14,578 qualified brain ICs), 77.6% of fitted dipoles lay more than 5 mm inward from the template boundary and 57.9% lay more than 14 mm inward. The pooled signed-distance distribution showed two prominent peaks, near −1 mm and −19 mm, separated by a pronounced minimum near −5 mm (2-mm bins). Fitted dipole orientations showed no tangential dominance among deep ICs, which also exhibited higher activation power and lower residual variance than superficial ICs. These results favor the large-patch model. ICA remains valid for separating EEG sources, but the depths of fitted dipoles may reflect the spatial extent of the underlying current sources.

## Introduction

This is a methods paper on the physiological interpretation of independent component analysis (ICA) (Bell & Sejnowski, 1995; Comon, 1994; Hyvärinen et al., 2001) applied to scalp EEG (hereafter, ICA means ICA application to EEG). However, to elucidate the historical and conceptual origins of this physiological interpretation, the scope of this paper extends beyond methodological research to encompass historical analysis, a literature review, and hypothesis-driven original research, with equal weight given to each. Accordingly, in the Introduction, before motivating the testable hypothesis, we first explain how the concept of the *small-patch model*, motivated by the microanatomy of intracortical lateral connections, became the central biophysical assumption underlying the conventional physiological interpretation of ICA. We then review the clinically and biophysically grounded alternative, the *large-patch model*, which has long existed outside the community of ICA users, and finally state the hypothesis and study design. A more detailed philological critique, quoting the original sources verbatim, is presented in the Discussion.

### Motivation of the current study

ICA is widely used to remove artifacts from EEG (Castellanos & Makarov, 2006; Chaumon et al., 2015; Delorme, Sejnowski, et al., 2007; Jung, Makeig, Humphries, et al., 2000; Jung, Makeig, Westerfield, et al., 2000; Mognon et al., 2011; Nolan et al., 2010; Winkler et al., 2011). Beyond artifact removal, ICA-derived scalp topographies are fitted with equivalent current dipoles to localize ICs to brain regions (Debener et al., 2005; Delorme, Westerfield, et al., 2007; Jung, Makeig, McKeown, et al., 2001; Jung, Makeig, Westerfield, et al., 2001; Makeig & Onton, 2011; Makeig et al., 2002; Makeig, Debener, et al., 2004; Makeig, Delorme, et al., 2004; Onton & Makeig, 2006; Onton et al., 2005, 2006). Although this use of ICA is less common, it has become an established practice and can be found in recent studies (Huang et al., 2026; Lahijanian et al., 2024; Phillips et al., 2023; Symeonidou & Ferris, 2023). This practice rests on an auxiliary assumption that “*there are strong biological reasons to believe that under favorable circumstances ICA should separate signals arising from local field activities in physically distinct, compact cortical source areas*” (Delorme et al., 2012) (p.1) in which “*expected effective size is relatively small (circa 1 cm or less)*” (Akalin Acar et al., 2016) (p. 171). However, this assumption is seldom explicitly confirmed and, to our knowledge, has not been directly tested. If ICs arise from synchronous patches spanning several gyral crowns, single dipole model fits to their scalp projections should be displaced systematically beneath the cortex (Fuchs et al., 2017; Scherg et al., 2019). Hereafter, we refer to this as *depth bias*. Consequently, IC selection based on fitted dipole locations relative to the gray matter (Jungnickel & Gramann, 2016; Lin et al., 2018) may reject ICs with apparently implausible source depth, even though such depths are what extended generators predict.

### Justification of ICA on EEG: independence-dipolarity identity (IDID)

ICA often yields near-dipolar scalp projections consistent with anatomically plausible cortical sources, despite using no spatial or biophysical constraints (Delorme et al., 2012; Onton & Makeig, 2006). In ERP studies, successful ICA decomposition has repeatedly been accompanied by scalp projections attributable to anatomically plausible sources (Debener et al., 2005; Delorme, Westerfield, et al., 2007; Makeig et al., 2002; Makeig, Delorme, et al., 2004; Makeig, Westerfield, Jung, et al., 1999; Makeig, Westerfield, Townsend, et al., 1999; Mouraux & Iannetti, 2009). Delorme et al. (2012) quantified this relation: across 18 ICA and related algorithms, the greater the reduction in mutual information between components, the larger the number of near-dipolar components. This correspondence between temporal independence and spatial dipolarity has been taken as evidence for the *physiological validity of ICA* (Delorme et al., 2012); we refer to it as the *independence–dipolarity identity (IDID)*.

This almost unreasonable effectiveness of ICA in EEG analysis may be explained by three properties of EEG generation. First, volume conduction mixes source activities linearly and instantaneously, and this process is modeled by a single matrix multiplication. However, this condition is shared by all linear decompositions and not unique to ICA. Second, volume conduction performs spatial averaging through linear mixing, which, by the central limit theorem, makes scalp observations closer to Gaussian than their sources. Mixing therefore leaves a margin that ICA can exploit (Hyvärinen et al., 2001), which is “rather beautiful motivation behind ICA” (Friston, 1998). Third, empirical evidence suggests that functionally distinct EEG sources exhibit approximately independent temporal dynamics and spatially distinct distributions, consistent with the assumptions underlying ICA. However, it is important to note that ICA makes no assumption about the size or shape of the sources. We next review the origin of the small-patch model.

### A historical review of the small-patch model

The application of ICA to EEG was first reported in 1996 (Makeig et al., 1996) and was developed further through the mid-2000s, including high-profile journal publications such as *PNAS* (Makeig et al., 1997) and *Science* (Makeig et al., 2002). The size of the cortical sources was initially loosely described, as “compact” (Makeig et al., 2002) or “cm²-scale” (Makeig, Debener, et al., 2004). Onton and Makeig (2006) attributed the small size of the sources to short-range neuronal connections and, to our knowledge for the first time, gave a spatial scale for them (500 μm). In 2012, Delorme and colleagues opened their paper by stating that “there are strong biological reasons” to expect ICA to separate “compact cortical source areas”, and argued that “short-range (< 100 μm) lateral connections between cortical neurons are vastly more dense than longer-range connections” (Delorme et al., 2012). In the same year, an animation illustrating the small-patch model was made publicly available (https://youtu.be/6HTaRSiFFBg; Figure 1A). Akalin Acar et al. (2016) developed Simultaneous tissue Conductivity And source Location Estimation (SCALE), adopting source “compactness” as an optimization objective alongside goodness of fit. They specified the expected size as “circa 1 cm or less” which, taken as a diameter, corresponds to an area of less than 1 cm^2^ (Akalin Acar et al., 2016). Thus, the small-patch model is motivated by the microanatomy of lateral neuronal connections and assumes “fully or partially coherent across a single small cortical domain or patch” (Akalin Acar et al., 2016). However, to our knowledge, the source-size assumption has not been directly tested.

**Figure 1.**
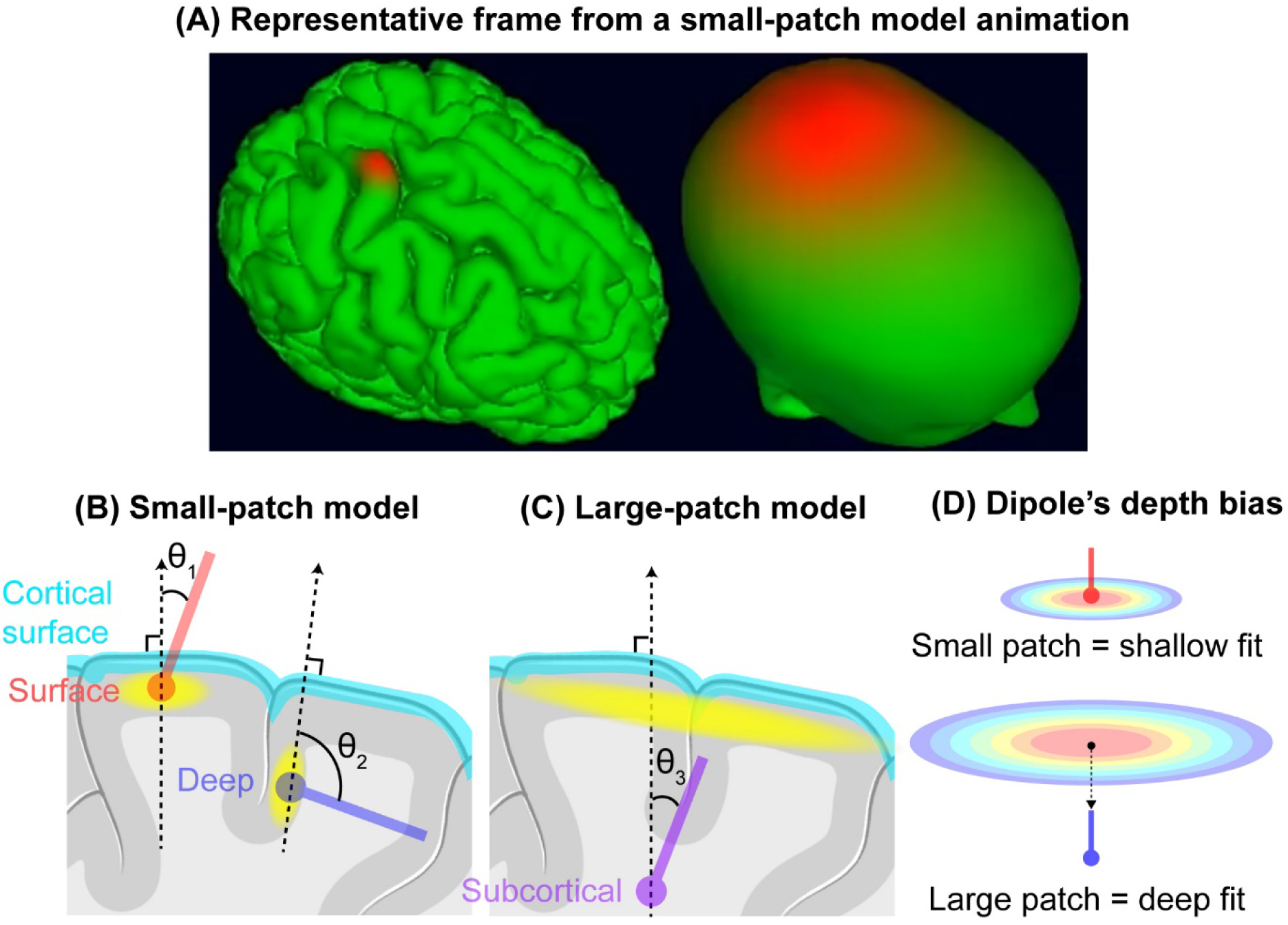
Illustration of the *small-patch* and *large-patch models* and their predicted effects on dipole depth and angles. **(A)** Screenshot from an animation of forward-model simulations generated using the Neuroelectromagnetic Forward Modeling Toolbox (NFT) (Akalin Acar & Makeig, 2010). The frame illustrates the scalp potential field produced by a localized cortical small-patch source. Image reproduced from a publicly available animation licensed under Creative Commons Attribution (CC BY https://www.youtube.com/watch?v=6HTaRSiFFBg published in 2012). (**B**) In the *small-patch model*, a synchronous cortical area smaller than a single gyral crown constitutes an EEG source. This model predicts that fitted equivalent current dipoles can distinguish gyral from sulcal sources by their depths and angles. Cortical patches highlighted in yellow represent main contributors to the scalp-recorded EEG signals. (C) In the large-patch model, by contrast, a synchronously active cortical region spans multiple gyri and sulci. The sulcal walls are active, but their contributions are substantially reduced by cancellation between opposing walls and by their greater depth. The net source is therefore the radial component from the gyral crowns. When such a broad source is approximated by a dipole, which is a spatially idealized point with no spatial extent, the fitted dipole is displaced deeper than the actual cortical layer (the *depth bias*). This model predicts that most estimated dipoles will be radially oriented and localized significantly deeper than the layer of the neocortex. (**D**) Schematic of dipole depth bias. An equivalent current dipole is a spatially idealized point source and provides a good approximation for a generator which is 3-4 times smaller than its distance to the electrodes (Nunez & Srinivasan, 2006). However, when a single dipole is used to approximate a potential field produced by a broad cortical patch, which is effectively a dense array of parallel dipoles aligned with the cortical normal, the fitted dipole is displaced to deeper locations. In this study, we refer to this systematic localization error as the *depth bias*, which we will later quantify in simulation.

### An alternative: the large-patch model

Outside the ICA community, the question of how large a synchronously active cortical area must be to produce a scalp-recordable EEG signal has been studied since the 1960s. A long-standing estimate places the minimum cortical area resolvable by scalp EEG at approximately 6 cm^2^ (i.e., 1 inch^2^) (Cooper et al., 1965). This minimum has been repeatedly replicated by multiple groups of epileptologists using simultaneous EEG-ECoG recordings (Cosandier-Rimélé et al., 2007; Ebersole, 1997, 2000; Hashiguchi et al., 2007; Ikeda et al., 1999; Kobayashi et al., 2005), and simulation studies have also supported its validity (Cosandier-Rimélé et al., 2007; Nunez & Srinivasan, 2006). Importantly, 6 cm^2^ represents a lower bound: examining inter-ictal spikes, Tao and colleagues found that 90% of cortical spikes arising from > 10 cm^2^ of activated cortex produced observable scalp potentials, whereas those arising from < 6 cm^2^ never did. They concluded that synchronous activation of 10–20 cm^2^ of gyral cortex is common (Tao et al., 2005, 2007), confirming the prediction from the simulation (Nunez & Srinivasan, 2006). We refer to the resulting view that scalp-recordable sources span at least 6–10 cm² of cortex as the *large-patch model* (Figure 1C). Throughout this paper, the two models are distinguished by area alone, and we read “compact” in the quoted literature as referring to size, not to spatial contiguity.

Why, then, do ICA-derived sources appear localized? Nunez and Srinivasan classified cortical sources of scalp EEG by spatial localization and temporal stationarity into four types: Type I, localized at fixed locations, such as lemniscal-mediated (Somervail et al., 2026) somatosensory evoked potentials (SEP) within 50 ms (Nunez & Srinivasan, 2006); Type II, localized at moving locations; Type III, distributed with a spatial pattern that is stable over time; and Type IV, distributed with a spatial pattern that changes over time. They argued that Type IV sources are “the rule rather than the exception” (Nunez & Srinivasan, 2006) (p. 195), and that localized sources are special cases. Ahlfors et al. (2010) similarly noted that “with the possible exception of early sensory evoked responses, patterns of activation are often sufficiently widespread that the effects of cancellation are relevant to interpretation of EEG and MEG data” (p. 145) (Ahlfors et al., 2010). The contrast can be quantified: the ratio of EEG to MEG peak-to-peak amplitude (μV to fT/cm) is about 0.04 for the median-nerve response at 20 ms, presumed to arise from a single focal dipole, but about 2.0 for sleep spindles, a 50-fold difference that was attributed to a distributed generator of EEG spindles (Dehghani et al., 2011). Although combining ICA with a single dipole model is well justified (Delorme et al., 2012), it may give the misleading impression that all ICA-derived EEG sources are Type I. This impression arises because the combination of ICA and single-dipole fitting reduces every source type to Type I in two steps. First, ICA removes spatial nonstationarity by fitting a time-invariant mixing matrix (Type II to Type I, Type IV to Type III). Second, the single equivalent dipole removes spatial extent by representing each IC scalp map by a point source (Type III to Type I). The central hypothesis of the current study is that the spatial extent removed in the second step does not vanish but reappears as depth bias.

Applied to cortical anatomy, the small-patch model makes a specific prediction. An average gyral crown is only 0.45–0.55 cm wide throughout adulthood (Díaz Caneja et al., 2021), so a patch about 1 cm across cannot be accommodated on most gyral crowns. In contrast, cortical sulci provide a substantially larger surface: with an average depth of 1.3–1.4 cm and a wall-to-wall width of 0.3–0.4 cm, a single sulcus provides roughly 3 cm of continuous cortical surface from shoulder to shoulder, and the average sulcal length is nearly 700 cm (Díaz-Caneja et al., 2021). Multiple 1 cm² patches could fit within this geometry. Consequently, the small-patch model predicts a strong dominance of sources arising from sulcal walls rather than gyral crowns.

### Competing predictions and study design

In this study, we test the two models against each other using ICA-derived equivalent current dipoles and show that the results support the large-patch model, thereby revising the assumptions about cortical source size underlying the physiological interpretation of ICA. As illustrated in Figure 1, the two models make distinct predictions regarding the depth and orientation of these dipoles. The predictions rest on three anatomical facts: only about one-third of the cortex lies on gyral crowns (Díaz-Caneja et al., 2021; Van Essen & Drury, 1997; Zilles et al., 1988), the average cortical thickness is approximately 2.5 mm, and the average sulcal depth is approximately 14 mm (Díaz-Caneja et al., 2021; Pakkenberg & Gundersen, 1997; Stark et al., 2007). If ICs sampled the cortex in proportion to its surface area, the small-patch model would predict that about one-third of brain ICs are localized to gyral crowns with small dipole angles relative to the radial direction, and about two-thirds to sulcal walls with large angles. Because a patch about 1 cm across cannot be accommodated on most gyral crowns, the sulcal fraction should be even larger. In either case, the fitted dipoles should remain within the cortical mantle, where the average sulcal depth is approximately 14 mm. The large-patch model instead predicts predominantly small dipole angles together with a general depth bias. Within a patch spanning several gyri, the opposing walls of each sulcus carry dipole layers of opposite orientation whose contributions largely cancel, leaving a net far field approximately radial to the cortical envelope (Ahlfors et al., 2010; Fuchs et al., 2017; Nunez & Srinivasan, 2006). When this broad field is fitted with a single equivalent dipole, volume conductor theory predicts that the fitted dipole is displaced deeper than the cortical layer, increasingly so as the patch area grows. Because this depth bias is known in principle (Ahlfors et al., 2010; Fuchs et al., 2017), we conducted a simple simulation with a four-layer spherical head model, parametrically varying dipole-layer area and depth, to calibrate its magnitude.

The two models also differ in the spatial structure of dipole orientations across the cortex and in how IC activation power should vary with depth (Table 1). Depth bias can also arise from an inaccurate head model, notably from misestimated skull conductivity (Akalin Acar et al., 2016). We therefore also test for a positive relation between IC activation power and dipole depth, which a global head-model error would not produce. To determine which model the data support, we analyzed a total of 820 EEG recordings (62 and 64 channels) from healthy adults during eyes-open and eyes-closed rest, drawn from two open datasets (Babayan et al., 2019; Getzmann et al., 2024). Table 1 summarizes the predictions of the two models.

**Table 1.** Comparison of predictions from the small-patch and large-patch models. Predicted observations are written in bold.

|  | Small-patch model<br>(anatomically realistic) | Large-patch model<br>(surface dominant) |
|---|---|---|
| Source locations | **70% of sources in sulci.**<br>*Surface*: on gyral crowns<br>*Deep*: on sulcal walls<br>*Subcortical*: (not predicted) | **All in the surface (gyral crowns).**<br>*Surface*: small continua of gyral crowns<br>*Deep*: large continua of gyral crowns<br>*Subcortical*: very large continua of gyral crowns |
| Var. in IC act. | **Follows inverse-square law.**<br>*Surface*: high IC activation power<br>*Deep*: low IC activation power<br>*Subcortical*: (not predicted) | **Increases with source area.**<br>*Surface*: low IC activation power<br>*Deep*: high IC activation power<br>*Subcortical*: very high IC activation power |
| Dipole angles | **Radial at gyrus, tangential at sulci.**<br>**Spatially high-freq distribution.**<br>**Mean at (0x1/3+90x2/3) = 60 degrees.**<br>Surface: radial<br>Deep: tangential<br>Subcortical: (not predicted) | **Always radial.**<br>**Spatially low-freq distribution.**<br>**Mean at 0 degrees.**<br>Surface: radial<br>Deep: radial<br>Subcortical: radial |

## Materials and Methods

### Simulation Study of Serial Forward-Inverse Source Modeling

#### Building a forward model

To demonstrate the depth bias i.e. the dependence of fitted single-dipole depth on the spatial extent of the true source, we conducted a simulation study using a 4-shell spherical head model. The radii of the cortex, cerebrospinal fluid (CSF), skull, and scalp shells were set to [7.9 8.0 8.5 9.0] cm. Lead fields were computed using the Fieldtrip function ft_compute_leadfield (Oostenveld et al., 2011). Sixty-four electrodes were used, with positions defined by the standard-10-5-cap385.elp montage distributed as a part of EEGLAB library. The electrodes used were Fp1, Fpz, Fp2, AF3, AF4, F7, F5, F3, F1, Fz, F2, F4, F6, F8, FT9, FT7, FC5, FC3, FC1, FCz, FC2, FC4, FC6, FT8, FT10, T7, C5, C3, C1, Cz, C2, C4, C6, T8, TP9, TP7, CP5, CP3, CP1, CPz, CP2, CP4, CP6, TP8, TP10, P7, P5, P3, P1, Pz, P2, P4, P6, P8, PO7, PO5, PO3, POz, PO4, PO6, PO8, O1, Oz, and O2. EEG sources were defined as uniform dipole layer (UDL) positioned 2 mm below the cortical surface. The radii of the UDL were parametrically varied (2, 4, 8, 16, 24, 32, 48, and 64 mm), corresponding to surface areas of 0.13, 0.50, 2.01, 8.04, 18.1, 32.2, 72.4, and 128.7 cm^2^, respectively. To account for uncertainty in skull conductivity, we simulated three brain-to-skull conductivity ratios (BSCR = 20, 40, and 80). Conductivity values for brain, CSF, skull, and scalp were [0.33, 1.65, 0.0165, 0.33] S/m for BSCR = 20 (Næss et al., 2017; Nunez & Srinivasan, 2006); skull conductivity was adjusted to 0.00825 and 0.004125 S/m for BSCR = 40 and 80, respectively. The UDL curvature matched that of the cortical shell. Forward-model scaling was expressed in µV/(nA·m)

#### Dipole fitting and sensitivity to skull conductivity mismatch

Dipole fitting was performed using FieldTrip (Oostenveld et al., 2011) as implemented in the EEGLAB DIPFIT plugin. A two-stage procedure was employed, consisting of an initial coarse grid search followed by a nonlinear gradient-descent refinement. The forward-inverse solution workflow is shown in Figure 3A. For ICs fitted with bilateral dipoles, the dipole with the larger moment magnitude was selected for the subsequent depth and orientation analyses.

To test whether errors in skull conductivity could account for or modify the depth bias of extended sources, we repeated the simulation with the BSCR varied independently in the forward and inverse models. All other parameters, including the conductivities of brain, CSF, and scalp, were unchanged from the original simulation (Figure 3C). For each forward BSCR (20, 40, 80) and UDL radius (1, 2, 4, 8, 16, 24, 32, 48, 64 mm), the scalp potentials were fitted with a single equivalent current dipole using each inverse BSCR, yielding 81 conditions (27 matched, 54 mismatched). The effect of mismatch was quantified as Δ*d* = *d*(*F, I, r*) – *d*(*F, F, r*), where *F* and *I* denote the BSCR values used in the forward and inverse models, respectively, and *r* is the UDL radius. Depth *d* was measured along the -axis from the brain surface at the vertex, with positive Δ*d* indicating deeper localization.

### Empirical Study of Large-Scale Resting-State EEG

#### Subjects

We analyzed EEG data from two publicly available datasets: the Leipzig Study for Mind–Body–Emotion Interaction (Leipzig dataset; n = 212 after preselection) (Babayan et al., 2019) and the Dortmund Vital Study (Dortmund dataset; n = 608) (Getzmann et al., 2024). For the Leipzig dataset, the final sample included 134 males; mean age = 39.3 years (SD = 20.3). Handedness distribution was 188 right-handed, 20 left-handed, and 4 ambidextrous participants. For the Dortmund dataset, the final sample included 232 males; mean age = 44.1 years (SD = 14.5). Handedness distribution was 566 right-handed, 40 left-handed, and 2 participants with missing handedness information. Some participants completed a 5-year follow-up measurement; however, follow-up sessions were not included in the present analysis.

#### Ethics statement

Data collection for both datasets complied with the Declaration of Helsinki. The Leipzig Study for Mind–Body–Emotion Interaction was approved by the ethics committee of the Medical Faculty of the University of Leipzig (reference number 154/13-ff). The Dortmund Vital Study was approved by the ethics committee of the Leibniz Research Centre for Working Environment and Human Factors (approval numbers A93-1 and A93-3 for the follow-up assessment). For both datasets, we confirmed upon download that all data were fully de-identified.

#### Task

Resting-state EEG with eyes open and closed was recorded in both datasets. In the Leipzig dataset, the recording consisted of 16 alternating 1-min blocks of eyes-closed and eyes-open conditions, totaling 16 minutes. In the Dortmund dataset, the resting-state session comprised a 3-min eyes-closed block followed by a 3-min eyes-open block, totaling 6 minutes.

#### EEG recordings

For the Leipzig dataset, scalp EEG was recorded using BrainAmp MR Plus amplifier with 62 active actiCAP electrodes (Brain Products GmbH, Munich, Germany) placed according to the international 10-10 system (Oostenveld & Praamstra, 2001): Fp1, Fp2, F7, F3, Fz, F4, F8, FC5, FC1, FC2, FC6, T7, C3, Cz, C4, T8, CP5, CP1, CP2, CP6, AFz, P7, P3, Pz, P4, P8, PO9, O1, Oz, O2, PO10, AF7, AF3, AF4, AF8, F5, F1, F2, F6, FT7, FC3, FC4, FT8, C5, C1, C2, C6, TP7, CP3, CPz, CP4, TP8, P5, P1, P2, P6, PO7, PO3, POz, PO4, PO8, and FCz. FCz served as the initial reference, which was later recovered at the cost of VEOG. EEG signals were recorded online with a band-pass filter of 0.015-1000 Hz at a sampling rate of 2500 Hz.

For the Dortmund dataset, scalp EEG was recorded using BrainAmp DC amplifier with 64 active actiCAP electrodes (Brain Products GmbH, Munich, Germany) placed according to the international 10-10 system: Fp1, Fp2, F7, F3, Fz, F4, F8, FC5, FC1, FC2, FC6, T7, C3, Cz, C4, T8, CP5, CP1, CP2, CP6, P7, P3, Pz, P4, P8, PO9, O1, Oz, O2, PO10, AF7, AF3, AF4, AF8, F5, F1, F2, F6, FT7, FC3, FC4, FT8, C5, C1, C2, C6, TP7, CP3, CPz, CP4, TP8, P5, P1, P2, P6, PO7, PO3, POz, PO4, PO8, TP9, TP10, FT9, and FT10. FCz served as the initial reference and was recovered by reducing the data rank by one (Kim et al., 2023). EEG signals were recorded online with a low-pass filter at 250 Hz and a sampling rate of 1000 Hz.

#### EEG preprocessing

Figure 2 provides an overview of the preprocessing pipeline. EEG preprocessing was performed using Matlab R2023a (The MathWorks Inc., Natick, MA) and EEGLAB library (Delorme & Makeig, 2004) with custom code (https://github.com/MakotoMiyakoshi/DS1). Raw EEG data were downsampled to 250 Hz. Canonical electrode locations on the MNI head template were assigned (Collins et al., 1994; Evans et al., 1993). A high-pass FIR filter (Hamming window; cutoff 1.5 Hz at -6dB; 1 Hz transition bandwidth) was applied. Artifact subspace reconstruction (ASR) was performed using the EEGLAB plugin clean_rawdata, with a cutoff threshold of SD = 20 (Anders et al., 2020; Chang et al., 2018, 2020; Kim et al., 2025; Kothe & Jung, 2016; Kothe & Makeig, 2013; Miyakoshi, 2023). No other options were used in applying clean_rawdata. EEG signals were then re-referenced to the average of all scalp electrodes plus the initial reference (continuous zero channel), thereby recovering FCz as the reference electrode (Kim et al., 2023). Adaptive mixture independent component analysis (AMICA) (Palmer et al., 2016) was applied, discarding data points exceeding 3 SD during the first 15 iterations (maximum 2000 iterations) only for the purpose of AMICA per developer’s recommendation (Jason Palmer, private communication). Independent components (ICs) were classified probabilistically using ICLabel (Pion-Tonachini et al., 2019) into the categories of brain, eye, muscle, heart, line noise, channel noise, and other. Finally, equivalent current dipole models were fitted to each IC scalp topography (columns of the ICA mixing matrix mapped to electrode locations) using FieldTrip (Oostenveld et al., 2011) and EEGLAB plugin fitTwoDipoles to fit symmetric bilateral dipoles where appropriate (Piazza et al., 2016).

**Figure 2.**
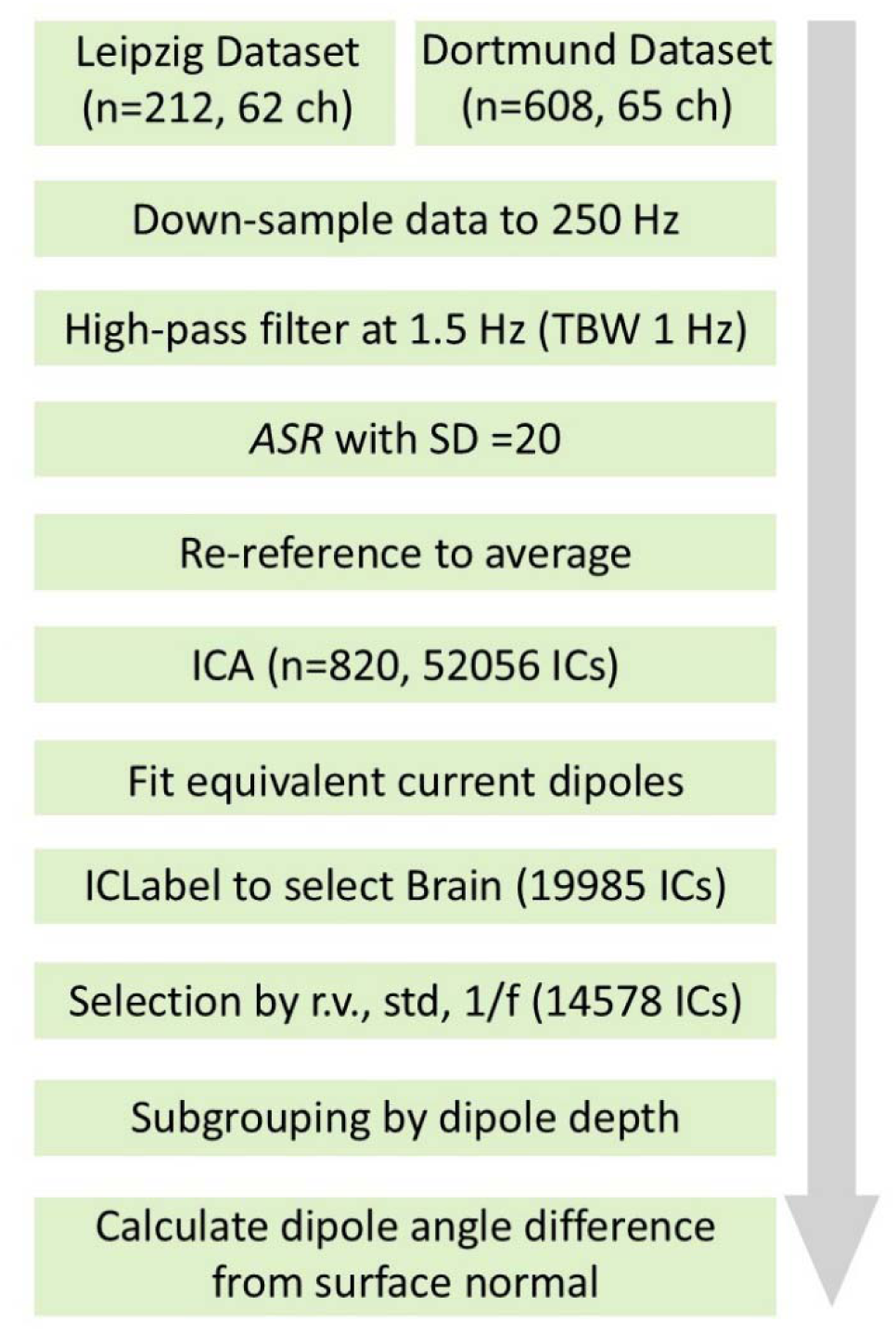
EEG preprocessing pipeline. ASR, artifact subspace reconstruction; ICA, independent component analysis; r.v., residual variance; std, standard deviation.

#### Head model preparation

For EEG source-model analysis, we used the ICBM152 template head model included with EEGLAB (avg152t1.mat). This anatomical template represents the averaged T1-weighted MRI of 152 healthy young adults (ages 18.5–43.5 years) acquired at the Montreal Neurological Institute as part of the International Consortium for Brain Mapping (ICBM) project (Fonov et al., 2011; Mazziotta et al., 1995)

Anatomical labeling was performed using the Automated Anatomical Labeling (AAL) atlas (Tzourio-Mazoyer et al., 2002) as implemented in the ROI_MNI_V4.nii file distributed with FieldTrip (Oostenveld et al., 2011). Because our analysis did not require internal compartment boundaries (e.g., ventricular walls or cerebellar interfaces), we manually edited lower axial slices (z = -26 to -12 mm) to close deep sulcal clefts and ventricular/cerebellar cavities, yielding a topologically spherical and approximately convex cortical volume. This customized cortical model was then expanded to match the spatial extent of the EEGLAB’s template head model ICBM152 on a slice-by-slice basis (see Figure S1 in Supplement). The resulting atlas was thus coregistered to the EEGLAB’s head model used for dipole fitting. The modified AAL cortical mask was used for anatomical classification. Dipole depth and orientation were calculated relative to a separate boundary mesh derived from the innermost (brain-compartment) surface of the three-layer EEGLAB DIPFIT template BEM head model (vol.bnd(3)). This surface was voxelized and processed to generate a closed mesh, which served as the common geometric reference for signed-distance and dipole-angle calculations. The resulting boundary is not equivalent to the individual cortical gray-matter surface.

For the primary angle analysis, we calculated polarity-invariant axial angles relative to the local outward normal at the nearest point on the template brain-compartment boundary. As a sensitivity analysis, we repeated the calculation using the radial direction from the MNI origin to each fitted dipole. To examine the dependence of the angle–depth relationship on the reference direction, we constructed joint histograms for both definitions using identical dipole populations and signed depths. Histograms were computed using fixed bin widths of 3° × 1 mm and smoothed with a two-dimensional Gaussian kernel (σ = 1.5 bins). For Figures 4–5, signed distance was defined as negative inward and positive outward. For Figure 8, dipole depth was defined as the negative of this signed distance, so that positive values indicate inward displacement. Axial angles were defined as *θ* = arccos (|*d* · *n*|), where *d* and *n* are unit vectors, yielding values between 0° and 90°. A separate set of 43 AAL labels was used as one criterion for identifying dipoles in the operationally defined Subcortex tier. These labels included the olfactory cortex, insula, hippocampus, parahippocampal gyrus, amygdala, caudate, putamen, pallidum, thalamus, and cerebellar regions. The tier name does not imply that all these structures are anatomically subcortical.

#### EEG analysis

Power spectral density (PSD) for IC activations was computed from 2 to 45 Hz in 0.1-Hz steps using MATLAB’s stft function with a 1-s Hamming window and 50% overlap. For robustness against outliers, the median across time windows was used instead of the mean. The spectral exponent coefficient *α* of the 1/*f^α^* spectrum was estimated using FOOOF (Donoghue et al., 2020) with the following parameters: aperiodic_mode = ‘fixed’, max_peaks = 1, min_peak_height = 0, peak_width_limits = [0.5 4], proximity_threshold = 0, peak_type = ‘best’, guess_weight = ‘weak’, hOT = 1, and thresh_after = true. The resulting exponent *α* from the 1/*f^α^* fit was later used for quality control. For each IC, the activation time series was divided into 100 nonoverlapping segments of equal length. The variance was calculated within each segment, and the median of these values was used as a robust estimate of IC activation variance. The ICA scalp projection maps were normalized to unit root-mean-square (RMS) amplitude across channels, ensuring a consistent scaling convention for comparing IC activation variances. Finally, residual variance from dipole fitting was also obtained for both quality control and evaluation.

ICs labeled as “brain” by ICLabel were further screened according to the following criteria:

1. residual variance of dipole fitting < 0.15
2. IC activation power falls between the 0.1 and 99.9 percentiles
3. spectral exponent coefficient α > 0.4. (Figure 4F)

Of the 14,796 ICs meeting the nonspatial quality criteria, 218 were excluded during spatial screening: 197 were fitted more than 10 mm outward from the template boundary, and 21 fell outside the anatomical atlas. The remaining 14,578 qualified Brain ICs were assigned to three operationally defined spatial tiers. Dipoles initially assigned outside the modified AAL cortical mask were reassigned to cortical candidates if they lay within 10 mm of the template boundary. Cortical candidates were divided into Surface cortex (less than 5 mm inward, including permitted outward locations) and Deep cortex (more than 5 mm inward). The Subcortex tier included noncortical candidates located more than 10 mm inward, dipoles identified by the selected AAL labels, and dipoles with MNI z ≤ −48 mm. This inferior-coordinate threshold was an operational classification criterion, not a subject-specific anatomical boundary. The AAL and inferior-coordinate criteria took precedence over the cortical classifications.

#### Interpretation of the AAL-defined cortex

It is important to note that the AAL-defined cortex is substantially thicker than actual human gray matter. Empirically, mean cortical thickness in the human brain is approximately 2.5 mm (Díaz-Caneja et al., 2021). The AAL cortical labels describe broad anatomical volumes and do not delineate cortical gray matter. In the modified mask, the mean thickness at z = 20 mm was 23.6 mm (SD = 6.2; range: 8.9–44.0 mm; Figure 4D), substantially exceeding cortical gray-matter thickness. These labels alone cannot identify dipoles fitted near cortical gray matter. We introduced an independent 5-mm inward-distance threshold to distinguish dipoles fitted near the template boundary from those fitted farther inward. This threshold is an operational criterion, not an anatomical estimate of cortical thickness. The resulting categories describe fitted equivalent-dipole locations, not the locations of their underlying current sources.

#### Software development and dissemination

For this study, we developed EEG_Clustering, an EEGLAB plugin extension for group-level analysis and visualization of spatiotemporal properties of ICs. The tool serves as an alternative to the EEGLAB STUDY framework by enabling IC clustering based on estimated dipole coordinates. It provides visualization of IC dipole-density distributions within the MNI template head space, as well as group-level summaries of associated time–frequency and spectral characteristics. The tool is available from https://github.com/kkukka21/EEG_Clustering and dedicated documentation is available in Supplement.

## Results

### Simulation Study of Serial Forward-Inverse Source Modeling

The forward-inverse simulation demonstrated depth bias in single-dipole fitting (Figure 3). As expected, this simple simulation revealed the area-depth dependency when using a single-dipole model. The overall residual variance was < 2% (for detail, see Supplement). We concluded that the large-patch model explains the depth bias when a single current dipole model is fitted.

**Figure 3.**
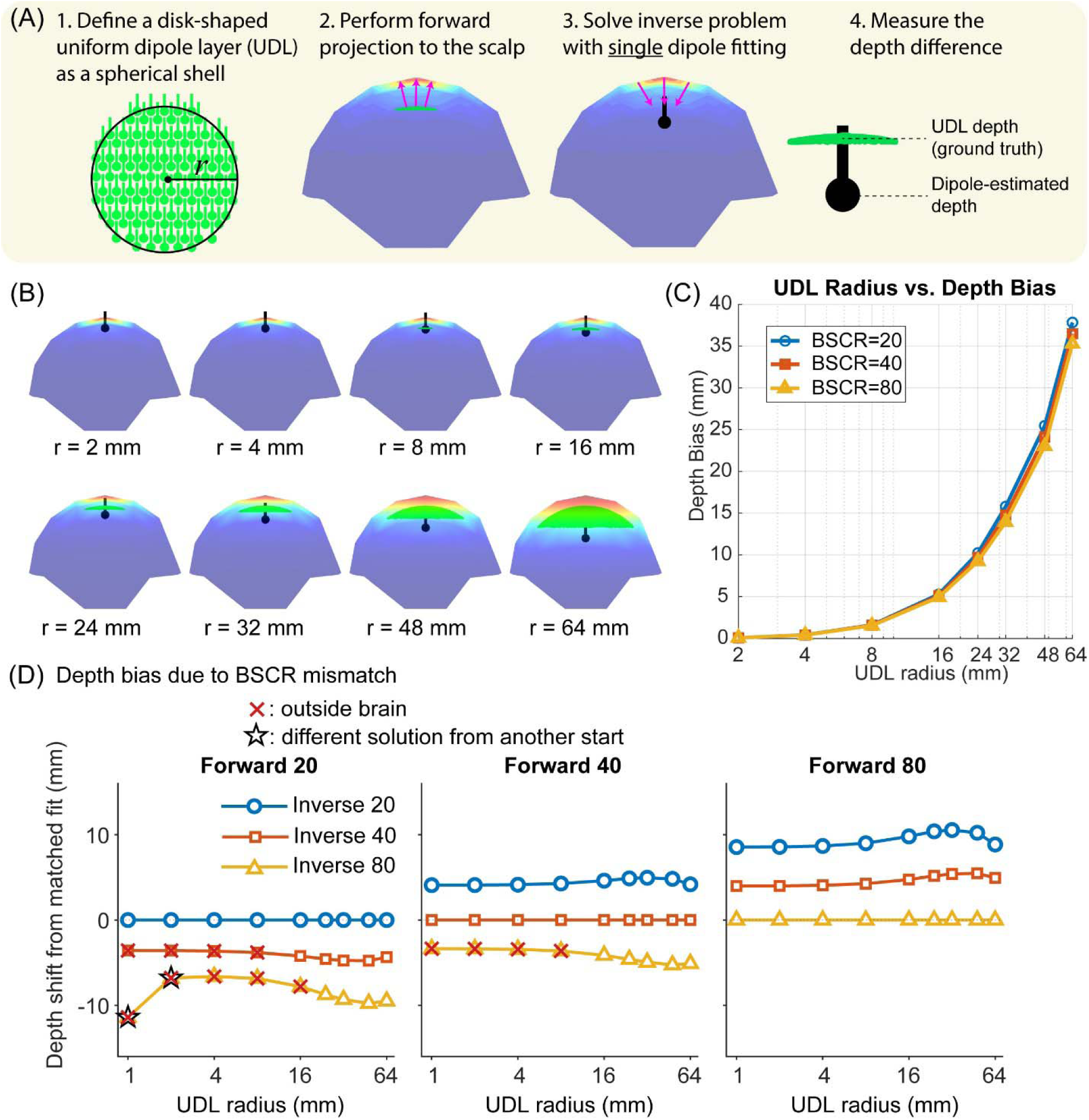
Forward-inverse simulation demonstrating depth bias in single-dipole fitting. A four-shell spherical head model was used with radii for the brain, cerebrospinal fluid (CSF), skull, and scalp set to [7.9 8.0 8.5 9.0] cm, respectively. Tissue conductivities were [0.33, 1.65, 0.0165, 0.33] S/m for the brain-to-skull conductivity ratio (BSCR) of 20; skull conductivity was adjusted to 0.00825 and 0.004125 S/m for BSCR values of 40 and 80, respectively. (**A**) Schematic of the simulation design. First, a disk-shaped uniform dipole layer (UDL), conforming to the curvature of the brain shell was defined with parametrically varied radii. The UDL depth was fixed at 2 mm below the brain surface and served as ground-truth source depth. Dipole orientations were set normal to the brain surface, with an inter-dipole spacing of 1.5 mm and unit dipole moments µV/(nA·m). Second, scalp potentials were generated via forward projection by computing lead fields from the UDL to 64 simulated scalp electrodes. Third, the resulting scalp topography was fitted using a single equivalent current dipole model. We predicted that broader source regions would yield systematically deeper estimated dipole locations. Finally, depth bias was quantified as the difference between the estimated dipole depth and the true UDL depth. In the schematic, the upper hemisphere rendered with a red-yellow-blue color scale represents the scalp potential distribution, the green convex disk represents the UDL, and the black disk-and-stem denotes the fitted dipole. For visual clarity, the UDL depth in (A) is exaggerated. (**B**) Estimated dipole locations for increasing UDL radii (r = 2–64 mm), demonstrating progressively deeper fitted dipoles despite constant source depth. (**C**) Radius-dependent depth bias across three BSCR conditions. Depth bias increases monotonically with UDL radius and is robust across skull conductivity assumptions. (**D**) Depth shifts caused by mismatch between the BSCRs assumed in the forward and inverse models. The three panels correspond to forward BSCRs of 20, 40, and 80, respectively; within each panel, the same simulated scalp-potential distribution was fitted using inverse BSCRs of 20, 40, and 80 across UDL radii of 1–64 mm. The vertical axis shows the change in estimated depth relative to the matched condition for the same forward simulation. Positive values indicate deeper estimates, and negative values indicate shallower estimates.

To assess the effect of skull-conductivity mismatch, we varied the BSCR independently in the forward and inverse models (Figure 3D). Because depth shifts are expressed relative to the matched fit, the zero lines for the matched conditions are definitional and do not indicate zero absolute localization error. Relative to the matched fits, a higher inverse BSCR shifted the dipoles toward the scalp, and a lower inverse BSCR shifted them deeper. With a forward BSCR of 80, inverse BSCRs of 40 and 20 increased the estimated depth by approximately 4–6 and 9–11 mm, respectively. With a forward BSCR of 20, inverse BSCRs of 40 and 80 decreased it by approximately 3–5 and 7–11 mm, respectively. A forward BSCR of 40 showed the same pattern. These shifts were nearly constant across UDL radii, whereas the matched depth bias grew with radius, reaching approximately 35–38 mm at 64 mm (Figure 3C). Fits outside the brain, and fits that depended on the initial dipole position, occurred only when the inverse BSCR exceeded the forward BSCR, and only for small radii (≤ 16 mm). Thus, skull-conductivity errors can amplify or partially offset the depth bias of extended sources, but within the tested range they did not eliminate its radius dependence.

### Empirical Study of Large-Scale Resting-State EEG

#### Extracting preselected brain components

Across the 820 datasets, we obtained 52,056 ICs, of which ICLabel identified 19,985 as Brain class candidates (Figure 4A). After applying our additional quality-control criteria (including the spectral-exponent criterion illustrated in Figure 4F), 14,578 ICs remained for the final analysis (Figure 4B). Class-wise analysis showed that excluded Brain-class ICs exhibited substantially lower activation variance than retained ICs, indicating their smaller contribution to the EEG variance (Figure 4E).

**Figure 4.**
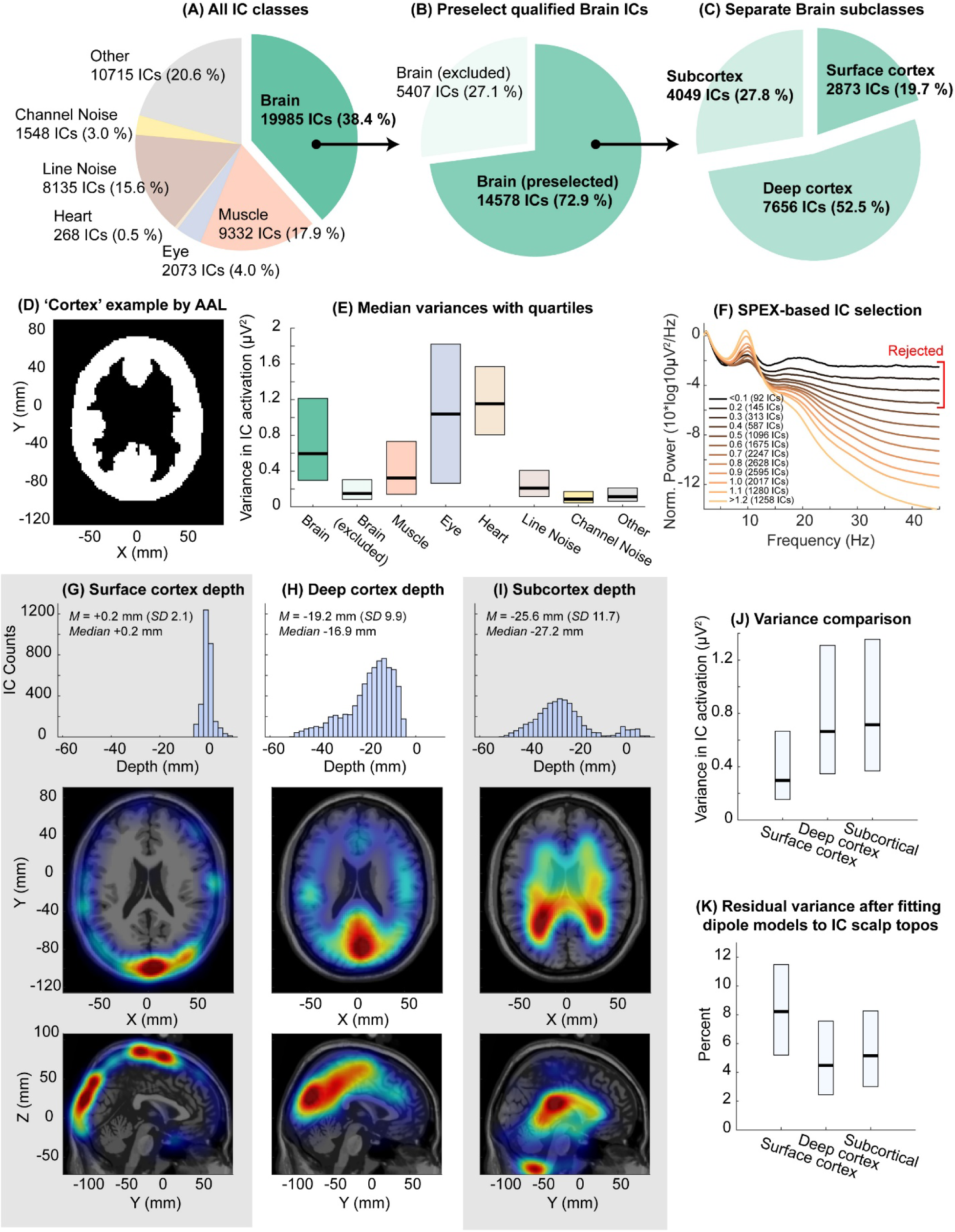
Selection of qualified Brain-class independent components (ICs) and summary statistics for spatiotemporal measures from 52,056 ICs across 820 EEG datasets. (**A**) Distribution of ICLabel classes. For each IC, the class with highest posterior probability was adopted. (**B**) After nonspatial quality screening, 14,796 ICs remained. Spatial screening excluded 197 dipoles located more than 10 mm outward from the template boundary and 21 outside the atlas, leaving 14,578 qualified Brain ICs. (**C**) The qualified Brain ICs were assigned to three operational spatial tiers: Surface cortex, Deep cortex, and Subcortex. Classification used distance from the template boundary, modified AAL anatomical labels, and an inferior-coordinate criterion, as detailed in Methods. (**D**) The modified AAL cortical mask at z = 20 mm had a mean thickness of 23.6 mm (SD = 6.2; range: 8.9–44.0 mm), substantially exceeding cortical gray-matter thickness. This illustrates the limitations of using volumetric AAL cortical labels as a proxy for gray matter. (**E**) Median IC activation variance with interquartile ranges for each ICLabel class. Qualified and excluded Brain-class ICs are displayed separately. (**F**) Mean power spectral density (PSD) across spectral-exponent (SPEX) bins, each spanning a 0.1 interval. ICs with SPEX < 0.4 were rejected because of their relatively flattened spectra. (**G-I**) Signed distances of fitted dipoles for the three operational tiers. Negative values indicate inward locations and positive values indicate outward locations. The displayed depth distributions overlap because the tiers were not defined exclusively by depth. Color limits (left to right panels): [0 0.05], [0 0.2], [0 0.1]. (**J**) Median and interquartile ranges of robust IC activation power for the three operational spatial tiers. (**K**) Median and interquartile ranges of residual variance after fitting dipole to IC scalp topographies. Smaller values indicate less errors.

#### Spatial classification and characterization of fitted dipoles

The retained Brain ICs were assigned to three operationally defined spatial tiers: Surface cortex (19.7%), Deep cortex (52.5%), and Subcortex (27.8%) (Figure 4C, G-I). The pooled signed-distance distribution showed two prominent peaks, near -1 mm and -19 mm, separated by a pronounced minimum near -5 mm (2-mm bins; Figure S7). The independently chosen 5-mm inward threshold therefore coincided approximately with a minimum in the empirical distribution. Overall, 77.6% (11,315 of 14,578) of qualified Brain ICs were fitted more than 5 mm inward from the template boundary. This proportion ranged from 74.9% to 79.6% when the threshold was varied from 3 to 7 mm.

Because sulcal walls extend well below gyral crowns, dipole depth alone cannot reliably distinguish the two models. Nevertheless, the depth distribution was strongly shifted inward: 57.9% (8,437 of 14,578) of qualified Brain ICs were fitted more than 14 mm inward from the template boundary. This distance is not directly equivalent to anatomical sulcal depth. Sulcal-wall generators should predominantly produce tangential dipoles with bipolar, figure-eight scalp topographies. However, the deep ICs showed no corresponding tangential dominance. Under both geometric reference definitions, the joint angle–depth distributions showed broad ranges ofdipole axial angles among deeply fitted ICs, without the concentration near 90° expected from predominantly tangential sulcal-wall generators (Figure 8). However, the apparent relationship between angle and depth differed substantially between the two reference directions.

A total of 4,049 ICs (27.8%) were assigned to the operational Subcortex tier. Of these, 3,615 met the noncortical distance criterion, 872 met the selected AAL-label criterion, and 314 met the inferior-coordinate criterion; these criteria overlapped. Thus, the Subcortex tier should not be interpreted as a purely anatomical classification. The predominance of inwardly displaced fitted dipoles, combined with the absence of the tangential dominance expected from sulcal-wall generators, is consistent with the large-patch model and difficult to reconcile with the small-patch model. Moreover, the combination of lower IC activation power and higher residual variance observed in Surface-cortex ICs indicates that these components represent a nondominant portion of the ICA decomposition and are generally of poorer quality (Miyakoshi et al., 2023). Conversely, ICs with higher activation power and lower residual variance were concentrated in the Deep-cortex and Subcortex tiers.

#### Depth tomography

Distribution of dipole depth within the brain space is visualized in Figure 5. It is noticeable that for axial slices in z >= 30 mm, the depth distribution is mostly concentric, while in 16 < z < 30 mm, a small island of low-depth region appears in the frontal region. A localized region of relatively shallow fitted dipole depths was observed in the frontal area at approximately z = 16– 30 mm. Its origin remains uncertain. Because depth was calculated relative to the template head-model boundary, this feature cannot be directly attributed to the internal anatomy or manually modified cavities of the AAL mask.

**Figure 5.**
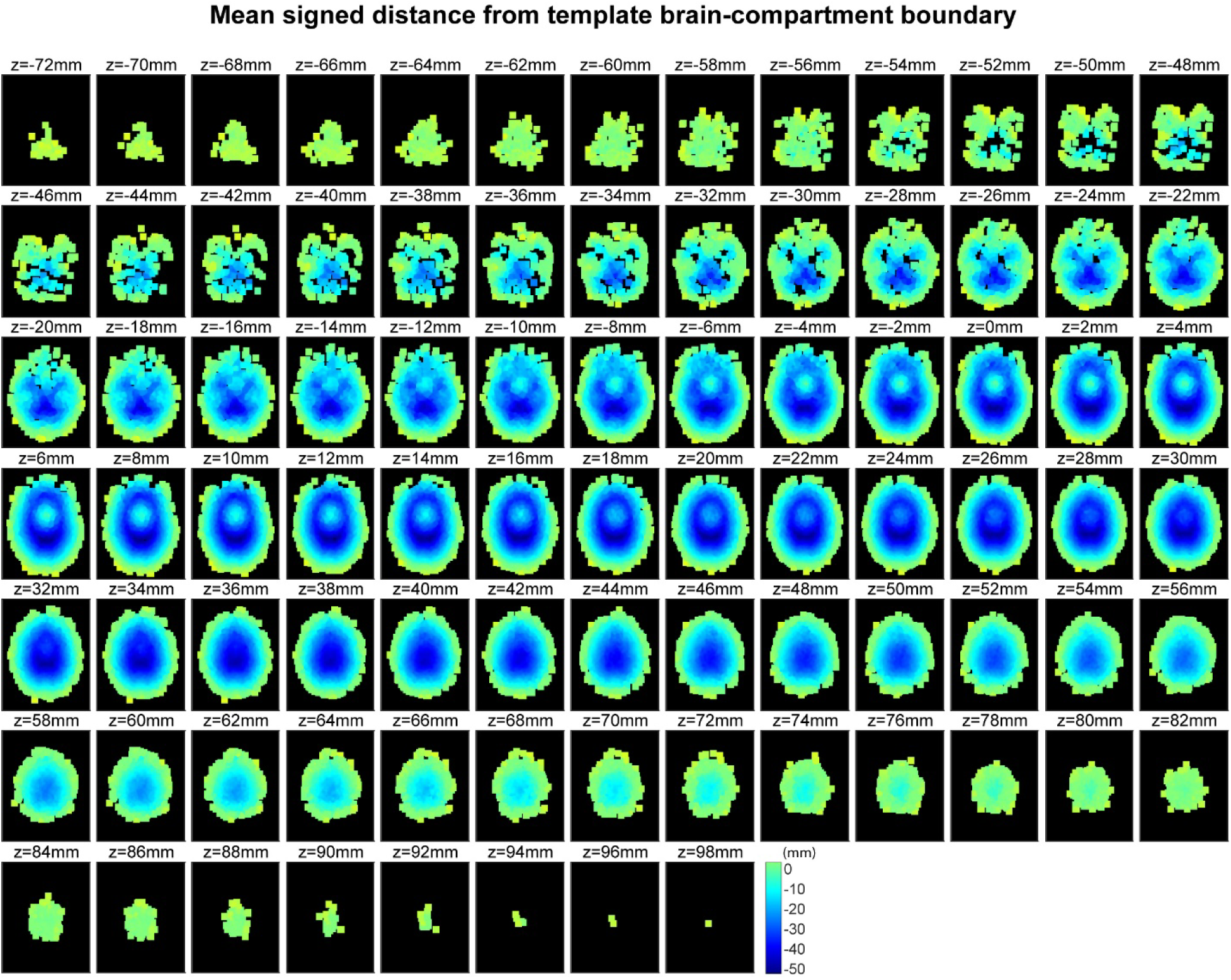
Mean signed distance of fitted dipoles relative to the template brain-compartment boundary, visualized across axial slices. Depth estimates were smoothed using a Gaussian kernel with σ = [2 2 2] mm.

#### Orientation tomography

The spatial distribution of dipole orientations across the brain is visualized in Figure 6. The small-patch model predicts that the orientation pattern should reflect the high spatial frequency of cortical gyrification. Given that the average gyral width is only 0.45–0.55 cm, which is approximately 3–4% of the 15–20 cm anterior–posterior brain axis, local cortical normals should rotate rapidly across the surface. Consequently, the small-patch model leads to a prediction of a highly heterogeneous, fine-grained, and nearly randomized pattern of dipole orientations throughout the brain volume. The empirical ICA-derived orientation tomography shown here however exhibits a clear contrast: broad, coherent, and low–spatial-frequency structure, which supports the large-patch model rather than the small-patch model.

**Figure 6.**
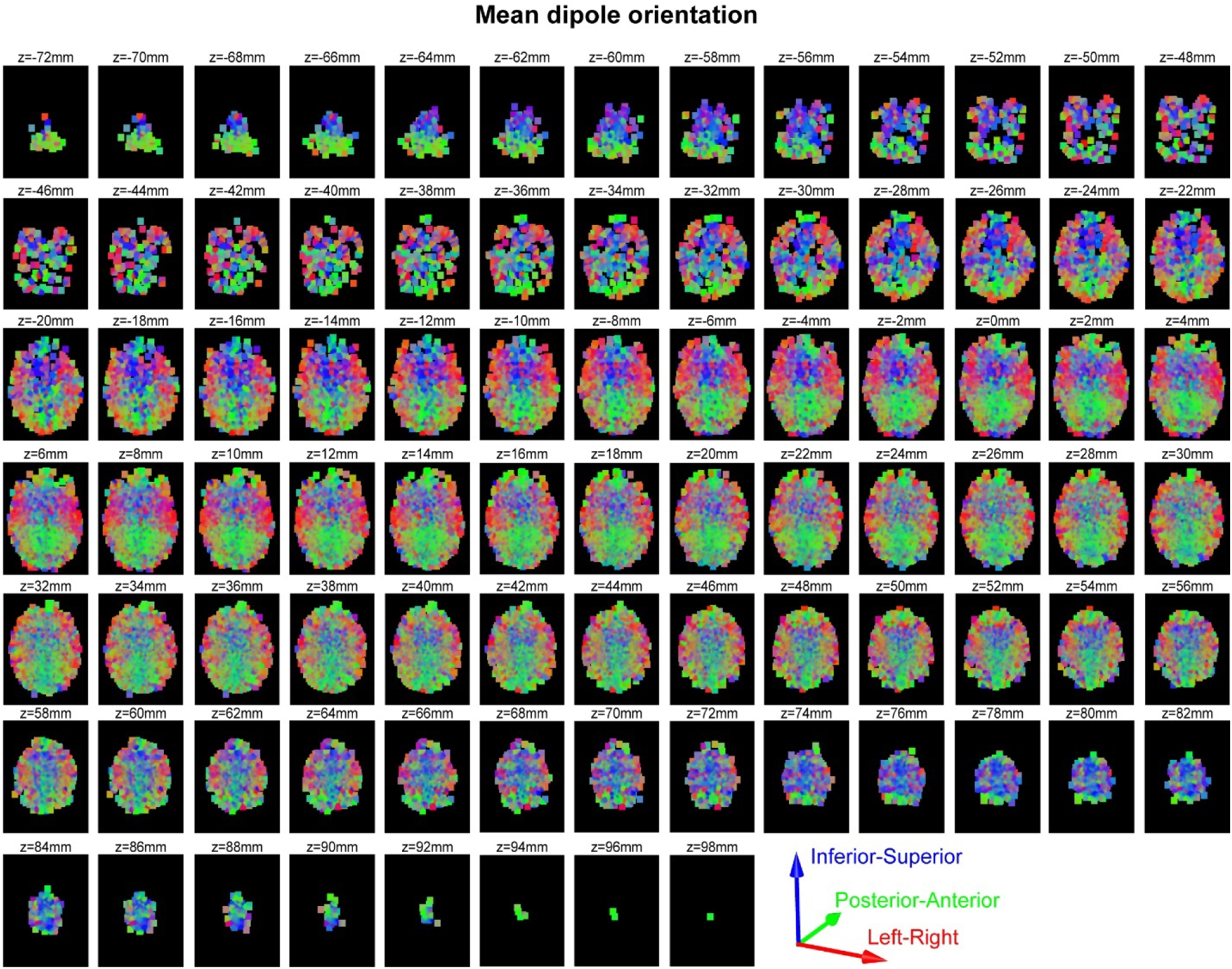
Mean dipole orientations, visualized across axial slices. The angles were smoothed using a Gaussian kernel with σ = [2 2 2] mm.

#### Angle-difference tomography, reference sensitivity, and angle–depth relations

The small-patch model predicts predominantly radial orientations at gyral crowns and tangential orientations along sulcal walls. However, a fitted dipole’s distance from the template brain-compartment boundary does not directly identify whether its underlying generator occupies a gyral crown or a sulcal wall. The empirical angle tomography in Figure 7A reveals broad spatial structure rather than an abrupt transition between radial and tangential orientations.

**Figure 7.**
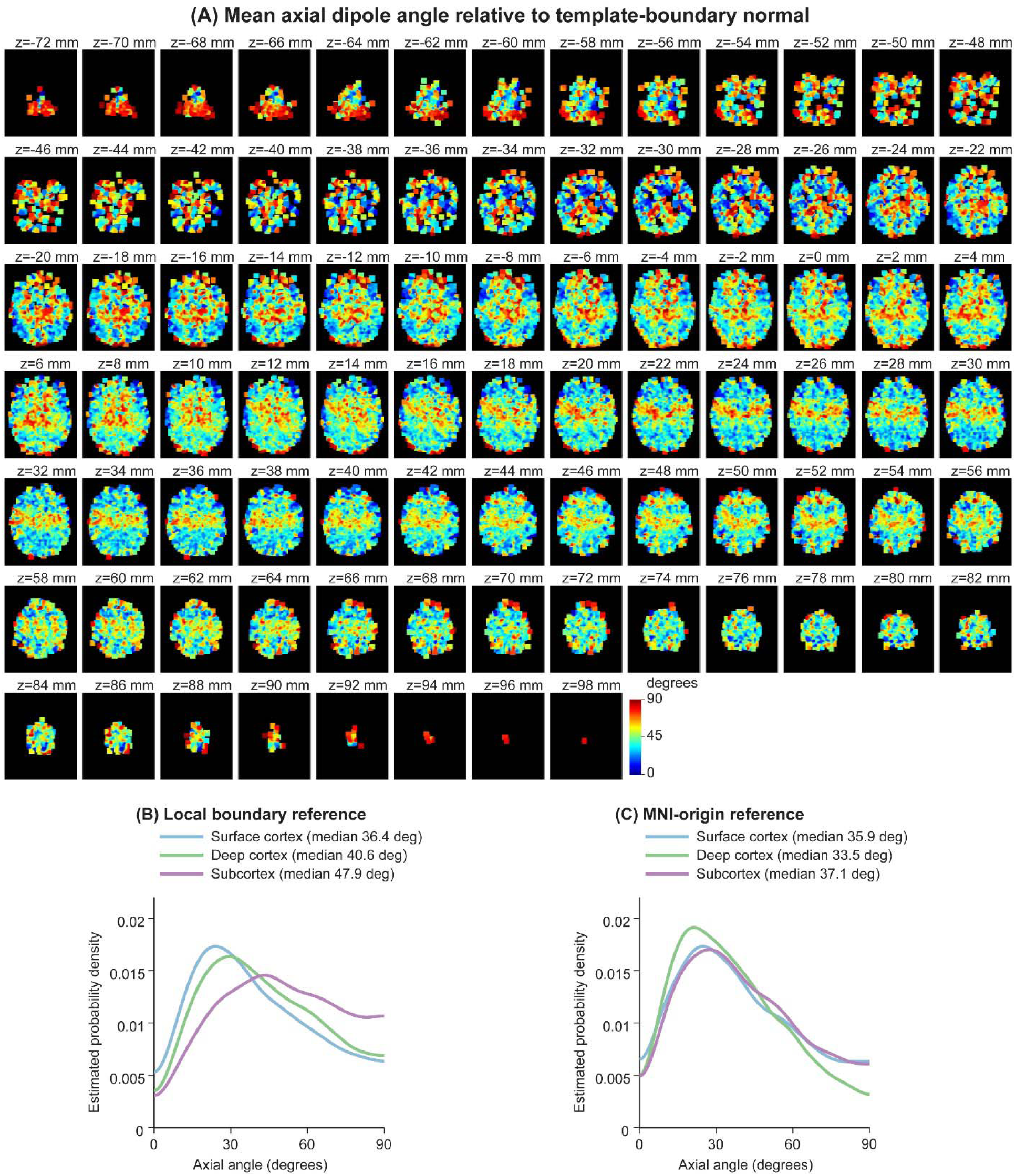
Spatial distribution of dipole axial angles and their dependence on the geometric reference direction. (A) Spatial distribution of mean dipole axial angles across 86 axial slices at 2-mm intervals (MNI z = −72 to 98 mm). For each dipole, the angle was calculated relative to the outward surface normal at the nearest point on the template brain-compartment boundary. Angles were spatially averaged using a Gaussian kernel (σ = 2 mm). Colors represent mean axial angles from 0° (radial, blue) to 90° (tangential, red). (B–C) Kernel density estimates of dipole axial angles for Surface cortex (n = 2,873), Deep cortex (n = 7,656), and Subcortex (n = 4,049), calculated relative to (B) the local template-boundary normal and (C) the radial direction from the MNI origin to the fitted dipole location. Angles were expressed as polarity-invariant axial angles (0–90°), and density estimates used Gaussian kernels with reflection boundary correction. Median angles for each group are indicated in the legends. All panels are based on 14,578 qualified Brain ICs.

A band of relatively larger axial angles was observed across approximately z = 16–56 mm (Figure 7A). Although this pattern may overlap peri-Rolandic regions, its precise anatomical origin cannot be determined from the present template-based tomography. Time–frequency signature in these regions is often characterized as the μ rhythm to which ICA is notably sensitive. This observation suggests that regional anatomical features may contribute to dipole-angle variability, although the relative contributions of anatomical curvature, fitted-dipole depth, and the choice of geometric reference direction cannot be determined from the tomography alone.

In contrast to the tangential dominance expected for sulcal-wall generators under the small-patch model, the local-reference distributions showed median axial angles of 36.4°, 40.6°, and 47.9° for Surface cortex, Deep cortex, and Subcortex, respectively (Figure 7B). None of the three distributions was concentrated near the tangential limit of 90°. This consistent dominance of moderately oblique orientations suggests the influence of additional, unresolved sources of angle variation at mesoscopic–macroscopic scales which were not captured by either the small-patch or large-patch idealizations.

To assess the sensitivity of dipole angles to the choice of reference direction, we compared two definitions: the local outward normal of the template brain-compartment boundary and the radial direction from the MNI origin to each fitted dipole (Figure 7B-C). To eliminate the arbitrary polarity of ICA-derived dipoles, angles were expressed as polarity-invariant axial angles ranging from 0° to 90°. The two definitions yielded nearly identical distributions for Surface cortex (median 36.4° vs. 35.9°), but increasingly different distributions for Deep cortex (40.6° vs. 33.5°) and Subcortex (47.9° vs. 37.1°). Thus, the estimated radiality of deep equivalent dipoles is sensitive to the geometric reference direction, whereas superficial dipoles are relatively insensitive to this choice.

To examine how the choice of geometric reference direction affects the apparent angle– depth relationship, we generated joint histograms using both the local boundary normal and the MNI-origin radial direction (Figure 8). Under the local-normal reference, peak angles increased from 22.5° for Surface cortex to 34.5° for Deep cortex and 40.5° for Subcortex (Figure 8A–C). In contrast, under the MNI-origin reference, the corresponding peaks were 25.5°, 22.5°, and 22.5° (Figure 8E–G), which replicated our previous report (Miyakoshi et al., 2023). Thus, the apparent increase in peak angle across depth tiers was not preserved when the reference direction was changed. Nevertheless, under both definitions, Surface-cortex ICs were concentrated near the template boundary, whereas Deep-cortex and Subcortex ICs occupied broadly overlapping regions of angle–depth space. These findings demonstrate the sensitivity of the apparent angle–depth relationship to the geometric reference direction. The reported peaks represent modes of smoothed joint histograms and do not directly quantify the conditional dependence of angle on depth. Taken together, the empirical distributions show no clear tangential dominance among deeply fitted dipoles under either reference definition. Although the local-normal histograms show increasing peak angles across depth tiers, this pattern is absent under the MNI-origin reference. Thus, the apparent angle–depth relationship is reference-dependent and cannot be interpreted as direct evidence of a transition from gyral to sulcal generators. These findings underscore the limitations of anatomical interpretations based solely on fitted equivalent-dipole geometry.

**Figure 8.**
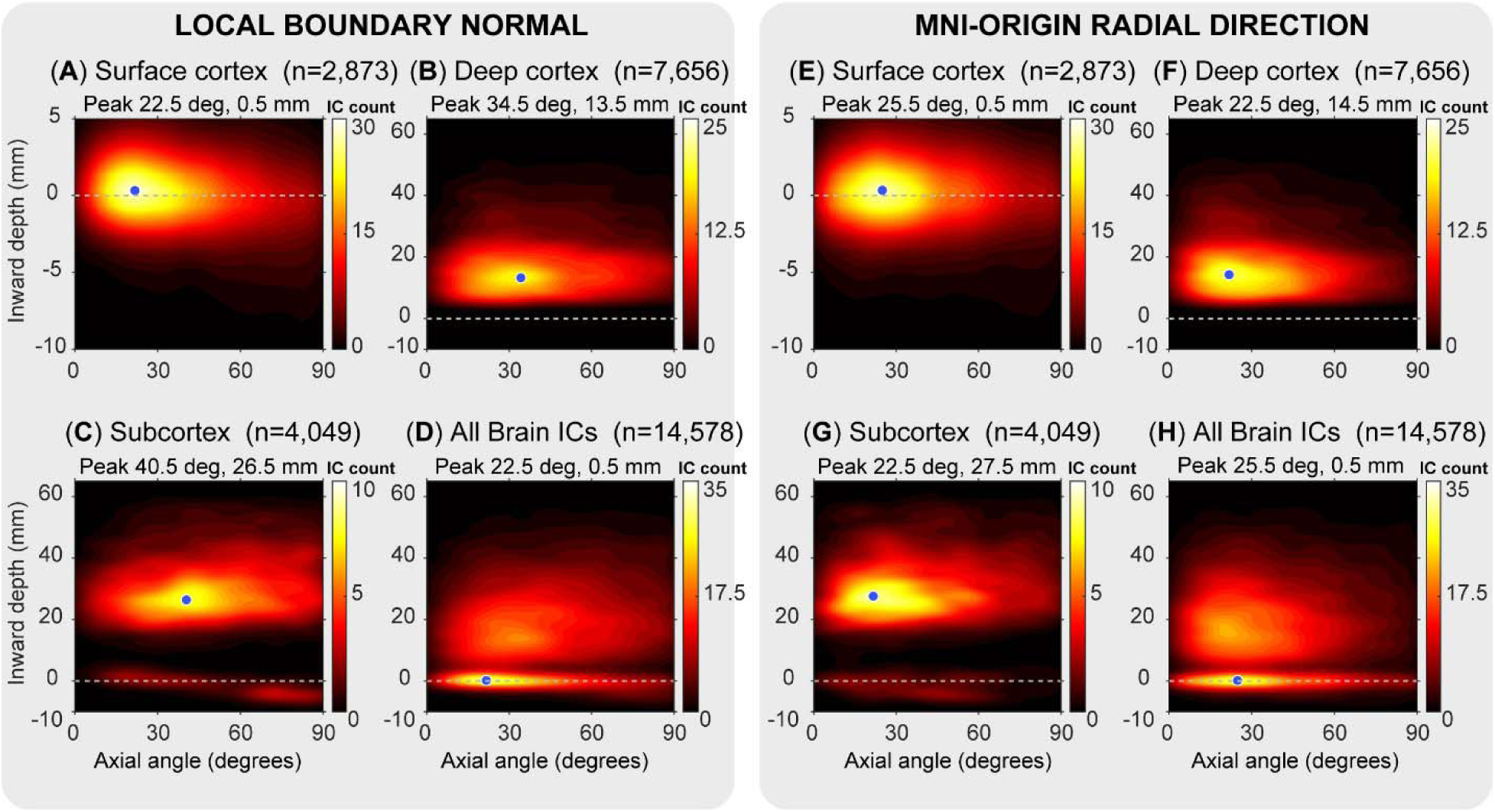
Joint histograms of fitted-dipole axial angle and depth using two reference directions. Joint distributions are shown relative to the local outward normal of the template brain-compartment boundary (left; **A**–**D**) and the radial direction from the MNI origin to the fitted dipole location (right; **E**–**H**). Each set includes Surface cortex (**A**, **E**; n = 2,873), Deep cortex (**B**, **F**; n = 7,656), Subcortex (**C**, **G**; n = 4,049), and all qualified Brain ICs combined (**D**, **H**; n = 14,578). Dipole orientations are expressed as polarity-invariant axial angles (0–90°), where 0° indicates alignment with the reference direction and 90° indicates perpendicular orientation. Dipole depth is the negative of the signed distance used in Figures 4–5, with positive values indicating inward displacement and negative values indicating outward displacement. Joint histograms were computed using fixed bin widths of 3° × 1 mm and smoothed with a two-dimensional Gaussian kernel (σ = 1.5 bins). Colors represent smoothed IC counts per bin, with matching color scales for corresponding panels across the two reference directions. Blue dots mark the peak-density bin centers, with their coordinates indicated above each panel. Dashed horizontal lines indicate the template boundary (0 mm). Panels **A** and **E** use an expanded depth scale to resolve the superficial distribution. The comparison illustrates how the apparent relationship between dipole orientation and depth depends on the choice of reference direction.

#### Validating the predictions of the small-patch model

Across all analyses reviewed above, Tables 2–5 summarize the extent to which the predictions of the small-patch model were supported or contradicted by the large-scale empirical database. In most comparisons, the observed distributions departed from the predictions of the small-patch model in dipole depth, orientation, or angle–depth structure. A narrow peak of fitted dipoles was observed near the template boundary (Figure 4G), separated from the broader inward peak by a pronounced minimum near −5 mm (Figure S7). This superficial peak is compatible with cortical generators near the boundary, although its precise location may be affected by head-model mismatch. Surface-cortex ICs showed lower activation power and higher dipole-fit residual variance than the deeper tiers. These differences indicate weaker components and poorer fits but do not exclude genuine superficial cortical generators. The principal challenge to the small-patch model is the predominance of deeper fitted dipoles without the expected tangential orientation pattern. Thus, ICs with stronger activation and better dipolar fits were frequently associated with unexpectedly deep equivalent-dipole locations. To our knowledge, this tension has not been discussed in the ICA literature.

**Table 2.** Small-patch model expectations and observed fitted-dipole locations. Signed distances are measured relative to the template boundary (negative inward, positive outward). The four distance intervals are mutually exclusive and collectively include all 14,578 qualified Brain ICs. Model expectations are qualitative where numerical predictions are unavailable. The 14-mm threshold is a descriptive reference to average sulcal depth, not an anatomical boundary.

| **Signed-distance interval** | **Small-patch model expectation** | **Observed ICs** |
|---|---|---|
| 0–10 mm outward | Possible due to localization errors | 1432 (9.8%) |
| 0–5 mm inward | Compatible with superficial cortical generators | 1831 (12.6%) |
| 5-14 mm inward | Compatible with sulcal-wall generators | 2878 (19.7%) |
| >14 mm inward | Possible for deep sulci or localization errors | 8437 (57.9%) |

**Table 3.** Small-patch model predictions and observed robust IC activation variance.

|  | IC activation power |  |
|---|---|---|
|  | Predicted by<br>Small-patch model<br>(inverse-square law) | Observed |
|---|---|---|
| Surface cortex | Large | *M* = 0.61 (*SD* 0.94), *Median* 0.30 |
| Deep cortex | Small | *M* = 1.12 (*SD* 1.50), *Median* 0.66 |
| Subcortical tier | No prediction | *M* = 1.20 (*SD* 1.67), *Median* 0.71 |

**Table 4.**
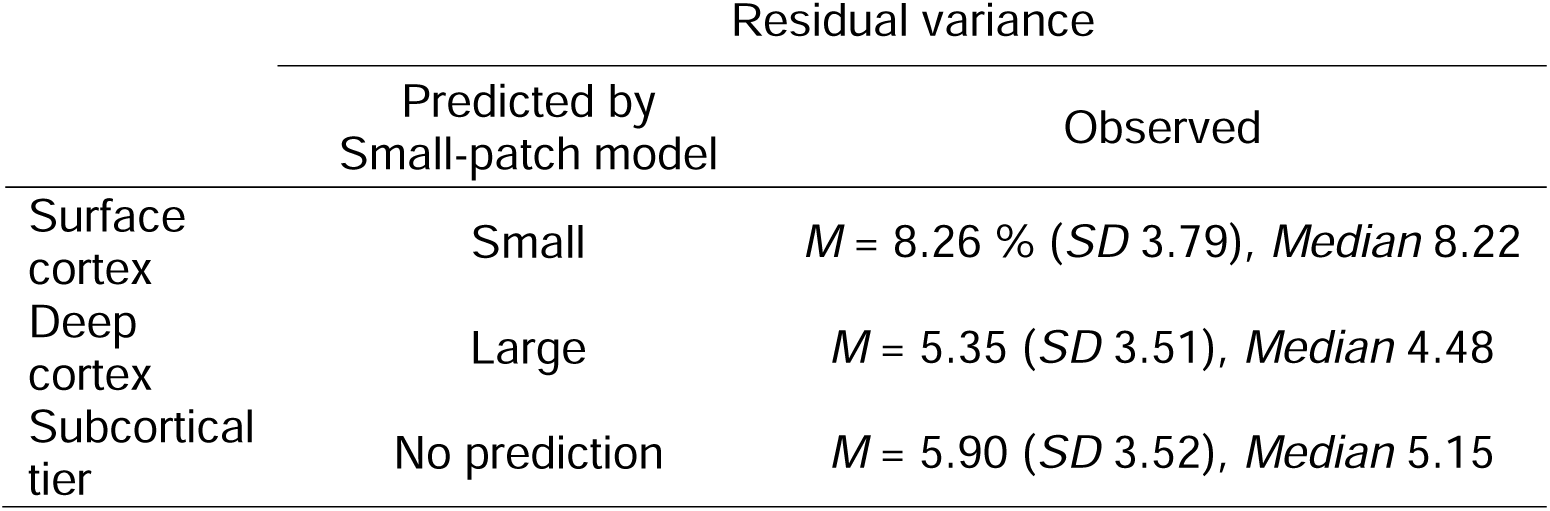
Small-patch model predicted versus observed residual variance after fitting dipole models to IC scalp topographies.

**Table 5.**
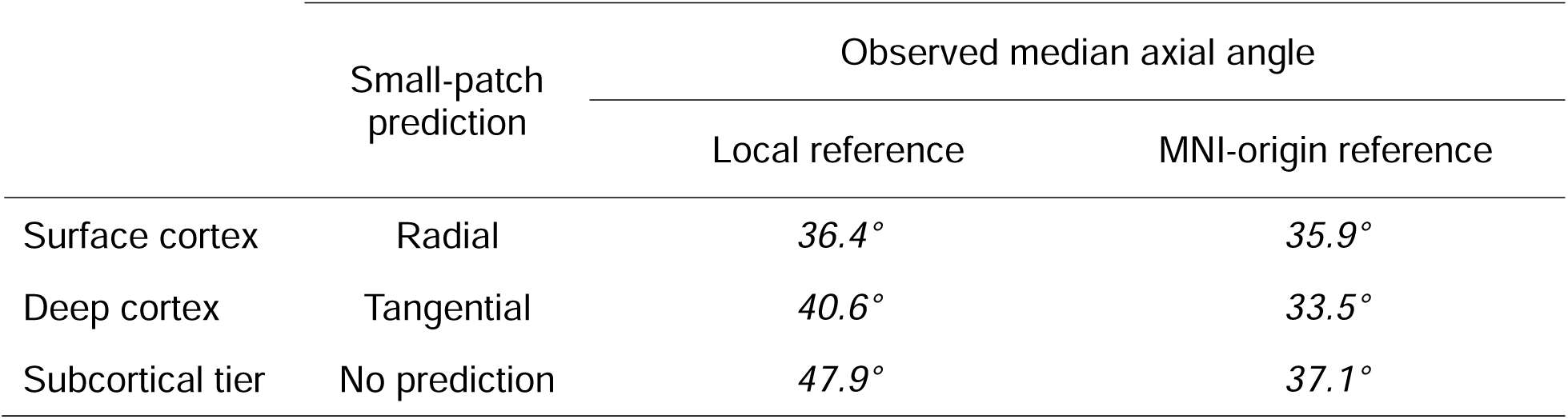
Small-patch model predictions and observed polarity-invariant fitted-dipole axial angles. Median values are reported separately for the local template-boundary and MNI-origin reference directions (Figure 7B–C).

## Discussion

In this study, we attempted to determine the physiological interpretation of ICA applied to scalp-recorded EEG. To this goal, we compared the small-patch source model proposed by early ICA proponents with the large-patch model supported by neurologists and theorists. Using two European resting-state EEG databases, we analyzed the distributions of ICA-derived dipole depth and their relationships to IC activation power, residual variance, and dipole orientation. Across these measurements, empirical results consistently favored the large-patch model over the small-patch model. Below, we discuss the implications of these findings point by point.

### The significance of the current study

The original physiological interpretation of ICA rests on the small-patch source model (Akalin Acar et al., 2016; Cao et al., 2012; Delorme et al., 2012; Makeig et al., 2002; Makeig, Debener, et al., 2004; Onton & Makeig, 2006; Tsai et al., 2014), an assumption that has not been directly tested, although simultaneous EEG–ECoG studies point to larger sources (Cosandier-Rimélé et al., 2007; Ebersole, 1997, 2000; Hashiguchi et al., 2007; Ikeda et al., 1999; Kobayashi et al., 2005; Nunez & Srinivasan, 2006; Tao et al., 2005, 2007). To our knowledge, the present study is the first to explicitly interrogate this core premise and test its predictions against large-scale empirical EEG data. Notably, despite the widespread adoption of ICA for artifact rejection, its use for extracting putative brain sources has remained limited. One plausible reason is that the physiological interpretation of ICA critically relies on dipolarity and the associated IDID (Delorme et al., 2012), yet ICA-derived dipole fits frequently yield physiologically implausible depths, which is a pattern supported by the predominance of inwardly fitted dipoles in the present data. Specifically, 77.6% of qualified Brain ICs were fitted more than 5 mm inward from the template boundary, and 57.9% more than 14 mm inward. Although these distances are not directly equivalent to anatomical sulcal depths, the observed depth distribution, together with dipole orientation, challenges the small-patch interpretation. Such results may have discouraged anatomical interpretation, in addition to the more technical challenge of post-ICA inter-subject inconsistency. A second reason concerns distributed source models. Dipolarity is not a physical quantity but a model-relative one: it is the goodness of fit of a single dipole. It therefore makes sense only within dipole fitting. Distributed source models applied to ICs provide no equivalent measure, so the validation performed by Delorme et al. (2012) cannot be carried out in that framework.

Distributed source modeling typically imposes a gray-matter constraint on source locations. In contrast, our dipole fitting does not impose any constraint on the location of fitted dipoles. Although the gray-matter constraint is physiologically motivated and computationally useful for solving the inverse problem, it carries the risk of prematurely discarding informative structure present in minimally constrained model outputs. The present study demonstrates the value of a minimally constrained model by examining outputs that may initially appear physiologically implausible. As shown in Figure 6 and 7, dipoles localized at implausible depths nevertheless exhibited systematic, spatially organized low-frequency patterns. This observation argues against dismissing deep dipoles. In fact, these deep dipole sources showed higher signal power and lower residual variance, indicating greater dominance and better model fit than superficially localized, physiologically plausible dipole sources. These observations caution against a bias toward premature physiological interpretation. Rather, judgment should be suspended (i.e., Husserlian *epoché*) until the full set of observations can be accounted for in light of basic anatomy and electrophysiology established in the literature. Finally, the presence of a systematic bias should not be regarded as bad as it may appear because it points to regularity in the underlying generative mechanism producing the error, like the case of the dipole’s depth bias we clarified using simulation.

Finally, the present study tested the small-patch model within the methodological framework of the original ICA studies. We used the same analysis tools and criteria as prior ICA-based work but examined two aspects that have received little attention: dipole depth and orientation. By applying the original approach to previously unexamined dimensions of the data, we test the small-patch model within its own methodological framework.

### Can head-model error explain the observed depths?

Skull-conductivity mismatch in the direction expected for our inverse model would shift fitted dipoles outward and therefore cannot explain the predominance of deep fitted locations. Our inverse model assumed a BSCR of 80, substantially higher than the value of 27.6 adopted in a recent account of EEG biophysics (Halnes et al., 2024). Although the actual BSCRs of our participants are unknown, this comparison suggests that the inverse model may have overestimated the ratio. As shown in Figure 3D, using an inverse BSCR higher than the forward BSCR shifts fitted dipoles toward the scalp. If the actual BSCR was below 80, this mismatch would oppose the inward bias associated with extended cortical generators. Our simulations also produced dipole estimates outside the brain boundary when the inverse BSCR exceeded the forward BSCR, particularly for small patches. In the empirical data, 1,195 of 2,873 Surface-cortex ICs (41.6%) were fitted outside the template boundary, with a median outward distance of 0.66 mm. This observation is consistent with the direction of the simulated conductivity-mismatch bias, although template anatomy, electrode-position errors, and other modeling uncertainties may also contribute.

### A canonical statement of the small-patch model

We selected a paragraph from Akalin Acar and colleagues (Akalin Acar et al., 2016) that we believe most clearly articulates the physiological concept underlying the small-patch model. It is a lengthy passage, but because it is central to the argument, we reproduce it in full below without modification.

More formally, we assume that each IC source represents a far-field projection to the electrodes of local cortical field potentials that are fully or partially coherent across a single small cortical domain or patch of unknown size and shape. In large part because of the extreme preponderance of short-range corticocortical connections (≤100 μm), wholly so for inhibitory neurons and glia, and the predominance of tight radial thalamocortical loops, both supporting oscillatory local field dynamics, models of cortical field dynamics (Deco et al., 2008) demonstrate the emergence of such patches or islands of cortical local field synchrony likened to “pond ripples” by Freeman (2003) and to recurring (point spread) avalanches by Beggs and Plenz (2004). Thus, brain connectivity both favors the emergence of temporal synchrony within small, connected cortical domains and minimizes temporal synchrony between such domains other than in exceptional locations and circumstances. Because of the common alignment of cortical pyramidal cells perpendicular to the cortical surface, spatially coherent local field activity across such a patch will have a non-negligible far field projection to the scalp (Baillet et al., 2001; Nunez and Srinivasan, 2006), forming an effective EEG source whose time course may typically be near statistically independent of the time courses of concurrent far field potentials generated by other, spatially separated cortical source patches.

Within this passage, the second sentence is of particular importance. At the same time, it is syntactically dense and conceptually layered. Focusing on this key sentence, we can simplify the argument as follows:

Because of the preponderance of short-range corticocortical connections (≤100 μm) and the predominance of radial thalamocortical loops, models of cortical field dynamics (Deco et al., 2008) demonstrate the emergence of small cortical patches.

In the following sections, we refer to this passage as the *canonical statement of the small-patch model,* and examine its validity.

### Where did the small-patch model come from, and what does it miss?

The small-patch model was motivated by electrophysiological studies conducted at microscopic spatial scales (≈100-1000 μm), including recordings from inhibitory (Budd & Kisvárday, 2001) and excitatory neurons (Stepanyants et al., 2009; Stettler et al., 2002) in the visual cortex of cats and macaques. These studies characterized the distribution of cortico–cortical lateral connection distances and emphasized the predominance of short-range connectivity such as ≤100 μm (Delorme et al., 2012) and < 500 μm (Onton & Makeig, 2006). From this microcircuit perspective, the original ICA studies inferred that EEG-measurable sources should arise from small cortical patches.

While this inference is intuitively appealing, it overlooks a critical property of population-level field generation that relatively small fractions of synchronized neurons can dominate macroscopic electrophysiological signals. Empirical and theoretical studies have shown that as little as 1% of pyramidal neurons within a cortical area of 1 cm² can account for approximately 96.5% of the observed signal amplitude (Hari, 1990; Hari et al., 1997). This counterintuitive effect arises from the nonlinear scaling of synchronous versus asynchronous contributions in large neuronal populations. To illustrate this point quantitatively, consider that the human cerebral cortex contains 22.9 billion neurons distributed across approximately 1,883 cm² of surface area in average man (Pakkenberg & Gundersen, 1997; Stark et al., 2007), with roughly 85% being pyramidal neurons (Kanari et al., 2019). This yields an estimate of approximately 10 million neurons per cm², which is remarkably similar to corresponding estimates in macaque visual cortex (O’Kusky & Colonnier, 1982). A synchronized subset comprising just 1% of this population corresponds to 100,000 neurons, whereas the expected amplitude of random, uncorrelated activity scales with the square root of the population size corresponds to 3162 neurons. Thus, 1% of synchronous neurons can produce 31.6 times larger signal than the background noise. The underlying simple mathematics shows that the contribution of a synchronized fraction always exceeds that of the entire population acting asynchronously once the number of neurons go beyond a critical mass 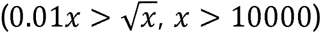, which is only modest in the spatial scale of EEG (Elul, 1971).

Independent empirical support for this sparse-synchrony, large-area view comes from the classic work of Walter (Walter et al., 1964). Using human intracranial EEG recordings, Walter observed that sensory-evoked responses rarely exceeded 30 µV, whereas direct electrical stimulation of the same cortical regions could elicit highly localized after-discharges of 1–3 mV, confined to only a few cubic millimeters of cortex. Interpreting these after-discharges as an analogue of “fully coherent” activation (as in the canonical statement) of the local neuronal population, Walter inferred that sensory-evoked responses engage on the order of 1% of neurons at any given cortical locus. The comparison on brain potentials between spontaneous field oscillation and that caused by electrical stimulation was replicated immediately (Cooper et al., 1965). Walter concluded that:

The conclusion is that a signal from any receptor, even if due to maximal sensory stimulation, can activate only about one neuron in a hundred in frontal cortex, but will have this effect over a vast surface containing several thousand million neurons. All significant sensory stimuli therefore […] must be considered as activating some tens of millions of neurons over a very extensive area, without necessarily intruding on any other afferent signals or background activity.

The scaling relationship reviewed above implies that scalp-recorded EEG is extremely sensitive to fluctuations in neural synchrony, and that this sensitivity grows rapidly with the spatial extent of the contributing cortical population. In contrast, the absolute synchrony rate becomes progressively less important as population size grows. By focusing on micro-scale connectivity and absolute synchrony rates, the small-patch model does not take into account this population-level property of field summation, thereby underestimating significance of the effective spatial extent of EEG-generating sources. Instead, scalp-recorded EEG is more plausibly dominated by weak neural synchrony distributed across large cortical patches (> 10 cm^2^ (Tao et al., 2005, 2007), corresponding to neural populations of > 100 million neurons (Kanari et al., 2019; O’Kusky & Colonnier, 1982; Pakkenberg & Gundersen, 1997; Stark et al., 2007)), with synchrony rate less than a few percent (Hari, 1990; Hari et al., 1997). This scaling law is both unique and critical to scalp EEG research, and fundamentally distinguishes it from invasive electrophysiological modalities.

Finally, we found that two anatomical claims commonly cited in support of the small-patch model do not match their sources. For example, in the canonical statement, work by Deco and colleagues is cited as evidence that an extreme preponderance of short-range corticocortical connections (≤100 μm) supports a small source model. However, Deco and colleagues explicitly stated that “Intracortical fibers are mostly unmyelinated and extend laterally up to 1 cm (in the human brain) with excitatory and inhibitory connections.” (Deco et al., 2008) Their work contains no analysis or claim linking the prevalence of ≤100-μm connections to the emergence of small patches of EEG sources. A second representative example appears in Delorme and colleagues (Delorme et al., 2012), who argued:

There are strong biological reasons to believe that under favorable circumstances ICA should separate signals arising from local field activities in physically distinct, compact cortical source areas: First, short-range (<100 μm) lateral connections between cortical neurons are vastly more dense than longer-range connections [3, 4]…

Reference [4] in this passage is the work of Stepanyants and colleagues (Stepanyants et al., 2009). However, Stepanyants and colleagues summarized their findings in the opposite direction: “excitatory connectivity in cat area 17 is highly nonlocal; 74% of excitatory synapses near the axis of a 1,000-μm-diameter cortical column originate from neurons located outside the column.” Thus, explicitly nonlocal connectivity (> 500 μm from the column axis) was cited as evidence for local connectivity (<100 μm). In both cases, the cited source does not support the claim attributed to it, yet these claims have been repeated for more than a decade and continue to reinforce the small-patch model. We suspect the reason lies in who reads which literature. The anatomical studies are read mainly by microcircuit physiologists, who have little occasion to open a methods paper on scalp-EEG ICA, and ICA users seldom go back to the primary anatomy. We have not surveyed the small-patch literature systematically and do not claim that these two cases are the only ones. Both discrepancies, however, are evident from the cited sources themselves, and readers should consult those sources before relying on anatomical claims passed along through this literature.

### Implications for the small-patch model

How much does challenging the small-patch model affect the physiological interpretation of ICA? We argue that the impact is limited because IDID (Delorme et al., 2012) is independent of the assumption of the source model and remains intact. That said, our findings have direct implications for Simultaneous Tissue Conductivity and Source Location Estimation (SCALE) (Akalin Acar et al., 2016). In this framework, skull conductivity is parametrically varied to identify a solution that minimizes the estimated cortical source patch size. While the methodological idea is innovative, the justification of the central optimization criterion for minimizing cortical patch size directly depends on the validity of the small-patch model. If the small-patch assumption does not hold, the physiological meaning of the SCALE solution becomes unclear, and the rationale for favoring minimal patch size over alternative models is no longer supported.

### Examples of a large-patch model

Whitmer and colleagues stated that “synchronous activity across a multi-cm cortical region has not been reported in the normal awake brain” (Whitmer et al., 2010). However, there are at least two counterexamples for this statement, which exemplifies the large-patch model across a multi-cm cortical region.

The first counterexample is the macroscopic traveling wave: simulation demonstrated that local coupling with neighbors supports globally extended spatiotemporal order (Jeong et al., 2002; Udeigwe & Ermentrout, 2015). Large spatial scales can be phase-organized without requiring long-range, millisecond-precise zero-lag synchrony. Synchrony is not a unitary phenomenon, and zero-lag synchrony across area is one extreme. The broad area can be coherent but not simultaneously. There are various phase gradients and travelling waves. The traveling wave phenomena in neural field has been known since early days of scalp EEG research (Hughes, 1995). There is a recent surge of publications on traveling waves from high-impact journals, with the emphasis on computational modeling (Dugué & Chavane, 2025) including (Halgren et al., 2019; Muller et al., 2018). In grey-matter recordings, Alexander and Dugué found that the spatial spectrum of cortical phase peaked at the longest measurable wavelengths (8–16 cm) from delta to high gamma, arguing against local generators blurred by volume conduction as the origin of scalp-recorded waves (Alexander & Dugué, 2026). These observations are difficult to reconcile with the small-patch premise that synchrony between cortical domains about 1 cm across is minimal (Akalin Acar et al., 2016): at the dominant wavelengths of 8–16 cm, sites 1 cm apart along the cortical sheet differ in phase by only about 20–45°.

Another counterexample shows more direct evidence of large-scale, simultaneous coherence mediated by extralemniscal thalamic pathway. It has been well-demonstrated that sudden and isolated sensory stimuli (SISS) trigger some of the largest and most widespread electrocortical responses in the awake mammalian brain mediated by the extralemniscal thalamic system: for review, see (Albe-Fessard & Besson, 1973; Miyakoshi et al., 2025; Somervail et al., 2026; Walter, 1964). It has some prominent characteristics including supramodality, sensitivity to inter-stimulus interval, vertex-maximum topography, and state of consciousness. In the current context, the most relevant and notable property of the extralemniscal thalamic system is that its projection is pan-cortical. The supramodality and pan-corticality was demonstrated using depth recording of mouse sensory cortex, in which auditory stimuli robustly triggered sensory cortex activation when inter-stimulus interval was 8 s (Benusiglio & Asari, 2024). Walter called non-modality-specific cortices and responses as non-specific cortex and response, respectively (Walter, 1964), the latter of which corresponds to responses of the extralemniscal thalamic system, and described it as follows:

Since the fields of the evoked potentials in nonspecific areas are so widespread that no part of the head is quite unaffected by them.

With this arrangement evoked responses to auditory, visual, and tactile stimuli have been recorded from the ears, chin, neck, and, in fact, all parts of the head.

The most satisfactory principle is to assume that, in experiments with human subjects, all evoked responses are mainly due to nonspecific activation unless the contrary can be proved.

To provide quantitative estimates, the ratio of thalamic neurons in the lateral geniculate nucleus to the cortical neurons they innervate is 0.1–1% (Connolly & Van Essen, 1984; O’Kusky & Colonnier, 1982). However, each individual axon is not restricted to a vertical column of these neurons, but spreads out tangentially and overlaps with its neighbors. As a result, this rate increases in the synaptic counts: The rate of cortex receiving thalamic input was classically estimated to be 5% in cat visual cortex (Peters & Payne, 1993), 6.9-8.7 % in macaque visual cortex (Latawiec et al., 2000). More recent estimate using a combination of 3D-electron-microscopy and 3D-confocal imaging of thalamic boutons on macaque visual cortex, 15 and 20% for magnocellular and parvocellular pathways, respectively, which is 2 to 3 times larger than the earlier estimates. (Garcia-Marin et al., 2019). In the rodent barrel cortex, the number has been around 15 % (Benshalom & White, 1986; Bruno & Sakmann, 2006; Schoonover et al., 2014). These quantitative estimates suggest that extralemniscal thalamo-pancortical projection contributing to form a large-patch cortical EEG source is anatomically plausible, contrary to the microanatomical argument of the small-patch model.

Simultaneous EEG–MEG recordings point the same way. Dehghani et al. (2011) proposed that EEG is most sensitive to sleep spindles generated diffusely and synchronously by the nonspecific (matrix) thalamocortical system, whereas MEG is most sensitive to multiple focal, asynchronous spindles generated by the specific (core) system (Dehghani et al., 2011; Somervail et al., 2026). This distinction parallels Walter’s separation of specific and nonspecific responses and suggests that scalp EEG preferentially records the extended activity driven by nonspecific thalamic projections. In Nunez and Srinivasan’s terms, the traveling wave is a Type IV source and the extralemniscal projection a Type III source. ICA reduces Type IV to Type III, and single-dipole fitting then converts the extent of either into depth bias; the present data therefore cannot distinguish them.

### Why do sulcal sources contribute little to dominant ICs?

How much do sulcal sources contribute to the dominant ICs of scalp EEG? About two-thirds of the cortical surface lies in sulci (Díaz-Caneja et al., 2021; Van Essen & Drury, 1997; Zilles et al., 1988), yet bipolar, 8-shaped topographies generated by tangentially oriented dipoles were rare among high-quality brain ICs. In our previous analysis of the same Leipzig data, positive-dominant ICs with low residual variance peaked at 10–30° from the radial axis, with no secondary peak near 90° (Miyakoshi et al., 2023). The geometry of an extended source explains this. When a patch spans a sulcus, the opposing walls carry dipole layers that face each other and point in opposite directions. Their far fields cancel to leading order, leaving a quadrupolar residual whose magnitude scales roughly with the ratio of the wall separation to the electrode distance. With the two grey-matter layers 5–7 mm apart and the electrodes 2–3 cm away, this residual is roughly 20–30% of the contribution of a single wall. The net field of a folded patch is therefore dominated by the radial contributions of the gyral crowns, which sum like a smooth dipole layer on the cortical envelope. The fundi are also radially oriented but contribute less because they occupy a smaller area and lie about 14 mm deeper, farther from the electrodes. (Ahlfors et al., 2010; Nunez & Srinivasan, 2006). Sulcal cortex is thus unlikely to form a dominant IC unless only one wall of a sulcus is active, in which case it appears as a tangential source. Such ICs are rare: we estimated them to account for at most about 9% of qualified brain ICs (Miyakoshi et al., 2023). The sensitivity analyses (Figures 7B–C and 8) further demonstrate that both the distribution of dipole angles and their apparent relationship with depth depend on the geometric reference direction. In particular, the increase in peak angle across spatial tiers observed under the local-normal reference was not reproduced under the MNI-origin reference. Neither reference necessarily corresponds to the orientation of local cortical columns.

### Conclusion

The present study showed that most ICA-derived brain components yielded deep fitted dipoles (77.6% more than 5 mm inward and 57.9% more than 14 mm inward from the template brain-compartment boundary), without the tangential dominance expected from the small-patch model. Moreover, the apparent relationship between dipole orientation and depth depended substantially on the geometric reference direction. Together, these findings challenge the conventional small-patch interpretation and favor the large-patch model as a more plausible alternative. A lesson from this work is that macroscopic EEG analysis requires a correct understanding of scaling laws: for example, how synchrony in as little as 1% of neurons can outweigh the square root of total but random activity. ICA itself remains valid for separating EEG sources; what changes is how the depths of fitted dipoles should be read. We hope that this study motivates future investigations into whether depth bias reflects a general property of EEG source modeling and how such biases may be explained by established and emerging biophysical theories.

## Data Availability

The EEG recordings analyzed in this study are publicly available from the Leipzig Mind-Brain-Body EEG dataset (LEMON; https://nemar.org/dataset/nm000179) and the Dortmund Vital Study (OpenNeuro accession ds005385; https://doi.org/10.18112/openneuro.ds005385). The MATLAB analysis code is available at https://github.com/MakotoMiyakoshi/DS1.

https://doi.org/10.18112/openneuro.ds005385

https://github.com/MakotoMiyakoshi/DS1

## Acknowledgement

MM thanks the following researchers for insightful discussions and contributions: Edmund Wascher for information about the Dortmund dataset; Hirokazu Tanaka for discussions on the independence–dipolarity identity of ICA; Robert Oostenveld for guidance on MATLAB simulation code and foundational physiological concepts; Paul Nunez for discussions on dynamic cortical source models; Giandomenico Iannetti for discussions on pan-cortical extralemniscal thalamic projections; and Scott Makeig for discussions on the small-patch model.

## CRediT author statement

**Komal K. Kukkar**: Methodology, Software, Formal analysis, Visualization, Writing – original draft.

**Hyeonseok Kim**: Methodology, Data curation, Writing – review & editing.

**Pranav J. Parikh**: Supervision, Project administration, Funding acquisition Writing – review & editing.

**Makoto Miyakoshi**: Conceptualization, Methodology, Software, Formal analysis, Investigation, Data curation, Visualization, Writing – original draft, Supervision, Project administration, Funding acquisition.

## Preregistration

This study and its analysis plan were not preregistered.

## Declaration of generative AI and AI-assisted technologies in the manuscript preparation process

During the preparation of this manuscript, the authors used Anthropic Claude and OpenAI ChatGPT to assist with manuscript editing, language refinement, and critical review. The authors reviewed and edited the resulting content and take full responsibility for the content of the published article.

## References

Ahlfors, S. P., Han, J., Lin, F.-H., Witzel, T., Belliveau, J. W., Hämäläinen, M. S., & Halgren, E. (2010). Cancellation of EEG and MEG signals generated by extended and distributed sources. Human Brain Mapping, 31(1), 140–149. 10.1002/hbm.20851

Akalin Acar, Z., Acar, C. E., & Makeig, S. (2016). Simultaneous head tissue conductivity and EEG source location estimation. Neuroimage, 124(Pt A), 168–180. 10.1016/j.neuroimage.2015.08.032

Akalin Acar, Z., & Makeig, S. (2010). Neuroelectromagnetic forward head modeling toolbox. Journal of Neuroscience Methods, 190(2), 258–270. 10.1016/j.jneumeth.2010.04.031

Albe-Fessard, D., & Besson, J. M. (1973). Convergent Thalamic and Cortical Projections — The Non-Specific System. In A. Iggo (Ed.), Somatosensory System (Vol. 2, pp. 489–560). Springer Berlin Heidelberg. 10.1007/978-3-642-65438-1_14

Alexander, D. M., & Dugué, L. (2026). The dominance of large-scale phase dynamics in human cortex, from delta to gamma. ELife, 13. 10.7554/eLife.100674

Anders, P., Müller, H., Skjæret-Maroni, N., Vereijken, B., & Baumeister, J. (2020). The influence of motor tasks and cut-off parameter selection on artifact subspace reconstruction in EEG recordings. Medical & Biological Engineering & Computing, 58(11), 2673–2683. 10.1007/s11517-020-02252-3

Babayan, A., Erbey, M., Kumral, D., Reinelt, J. D., Reiter, A. M. F., Röbbig, J., Schaare, H. L., Uhlig, M., Anwander, A., Bazin, P.-L., Horstmann, A., Lampe, L., Nikulin, V. V., Okon-Singer, H., Preusser, S., Pampel, A., Rohr, C. S., Sacher, J., Thöne-Otto, A., … Villringer, A. (2019). A mind-brain-body dataset of MRI, EEG, cognition, emotion, and peripheral physiology in young and old adults. Scientific Data, 6, 180308. 10.1038/sdata.2018.308

Bell, A. J., & Sejnowski, T. J. (1995). An information-maximization approach to blind separation and blind deconvolution. Neural Computation, 7(6), 1129–1159. 10.1162/neco.1995.7.6.1129

Benshalom, G., & White, E. L. (1986). Quantification of thalamocortical synapses with spiny stellate neurons in layer IV of mouse somatosensory cortex. The Journal of Comparative Neurology, 253(3), 303–314. 10.1002/cne.902530303

Benusiglio, D., & Asari, H. (2024). Sudden sensory events trigger modality-independent responses across layers in the mouse neocortex. BioRxiv. 10.1101/2024.11.07.622472

Bruno, R. M., & Sakmann, B. (2006). Cortex is driven by weak but synchronously active thalamocortical synapses. Science, 312(5780), 1622–1627. 10.1126/science.1124593

Budd, J. M., & Kisvárday, Z. F. (2001). Local lateral connectivity of inhibitory clutch cells in layer 4 of cat visual cortex (area 17). Experimental Brain Research, 140(2), 245–250. 10.1007/s002210100817

Cao, C., Akalin Acar, Z., Kreutz-Delgado, K., & Makeig, S. (2012). A physiologically motivated sparse, compact, and smooth (SCS) approach to EEG source localization. Annual International Conference of the IEEE Engineering in Medicine and Biology Society. IEEE Engineering in Medicine and Biology Society. Annual International Conference, 2012, 1546–1549. 10.1109/EMBC.2012.6346237

Castellanos, N. P., & Makarov, V. A. (2006). Recovering EEG brain signals: artifact suppression with wavelet enhanced independent component analysis. Journal of Neuroscience Methods, 158(2), 300–312. 10.1016/j.jneumeth.2006.05.033

Chang, C.-Y., Hsu, S.-H., Pion-Tonachini, L., & Jung, T.-P. (2018). Evaluation of artifact subspace reconstruction for automatic EEG artifact removal. Annual International Conference of the IEEE Engineering in Medicine and Biology Society. IEEE Engineering in Medicine and Biology Society. Annual International Conference, 2018, 1242–1245. 10.1109/EMBC.2018.8512547

Chang, C.-Y., Hsu, S.-H., Pion-Tonachini, L., & Jung, T.-P. (2020). Evaluation of Artifact Subspace Reconstruction for Automatic Artifact Components Removal in Multi-Channel EEG Recordings. IEEE Transactions on Bio-Medical Engineering, 67(4), 1114–1121. 10.1109/TBME.2019.2930186

Chaumon, M., Bishop, D. V. M., & Busch, N. A. (2015). A practical guide to the selection of independent components of the electroencephalogram for artifact correction. Journal of Neuroscience Methods, 250, 47–63. 10.1016/j.jneumeth.2015.02.025

Collins, D. L., Neelin, P., Peters, T. M., & Evans, A. C. (1994). Automatic 3D intersubject registration of MR volumetric data in standardized Talairach space. Journal of Computer Assisted Tomography, 18(2), 192–205. 10.1097/00004728-199403000-00005

Comon, P. (1994). Independent component analysis, A new concept? Signal Processing, 36(3), 287–314. 10.1016/0165-1684(94)90029-9

Connolly, M., & Van Essen, D. (1984). The representation of the visual field in parvicellular and magnocellular layers of the lateral geniculate nucleus in the macaque monkey. The Journal of Comparative Neurology, 226(4), 544–564. 10.1002/cne.902260408

Cooper, R., Winter, A. L., Crow, H. J., & Walter, W. G. (1965). Comparison of subcortical, cortical and scalp activity using chronically indwelling electrodes in man. Electroencephalography and Clinical Neurophysiology, 18, 217–228. 10.1016/0013-4694(65)90088-X

Cosandier-Rimélé, D., Badier, J. M., Chauvel, P., & Wendling, F. (2007). Modeling and interpretation of scalp-EEG and depth-EEG signals during interictal activity. Conference Proceedings: Annual International Conference of the IEEE Engineering in Medicine and Biology Society, 2007, 4277–4280. 10.1109/IEMBS.2007.4353281

Debener, S., Makeig, S., Delorme, A., & Engel, A. K. (2005). What is novel in the novelty oddball paradigm? Functional significance of the novelty P3 event-related potential as revealed by independent component analysis. Brain Research. Cognitive Brain Research, 22(3), 309–321. 10.1016/j.cogbrainres.2004.09.006

Deco, G., Jirsa, V. K., Robinson, P. A., Breakspear, M., & Friston, K. (2008). The dynamic brain: from spiking neurons to neural masses and cortical fields. PLoS Computational Biology, 4(8), e1000092. 10.1371/journal.pcbi.1000092

Dehghani, N., Cash, S. S., & Halgren, E. (2011). Emergence of synchronous EEG spindles from asynchronous MEG spindles. Human Brain Mapping, 32(12), 2217– 2227. 10.1002/hbm.21183

Delorme, A., & Makeig, S. (2004). EEGLAB: an open source toolbox for analysis of single-trial EEG dynamics including independent component analysis. Journal of Neuroscience Methods, 134(1), 9–21. 10.1016/j.jneumeth.2003.10.009

Delorme, A., Palmer, J., Onton, J., Oostenveld, R., & Makeig, S. (2012). Independent EEG sources are dipolar. Plos One, 7(2), e30135. 10.1371/journal.pone.0030135

Delorme, A., Sejnowski, T., & Makeig, S. (2007). Enhanced detection of artifacts in EEG data using higher-order statistics and independent component analysis. Neuroimage, 34(4), 1443–1449. 10.1016/j.neuroimage.2006.11.004

Delorme, A., Westerfield, M., & Makeig, S. (2007). Medial prefrontal theta bursts precede rapid motor responses during visual selective attention. The Journal of Neuroscience, 27(44), 11949–11959. 10.1523/JNEUROSCI.3477-07.2007

Díaz-Caneja, C. M., Alloza, C., Gordaliza, P. M., Fernández-Pena, A., de Hoyos, L., Santonja, J., Buimer, E. E. L., van Haren, N. E. M., Cahn, W., Arango, C., Kahn, R. S., Hulshoff Pol, H. E., Schnack, H. G., & Janssen, J. (2021). Sex differences in lifespan trajectories and variability of human sulcal and gyral morphology. Cerebral Cortex, 31(11), 5107–5120. 10.1093/cercor/bhab145

Donoghue, T., Haller, M., Peterson, E. J., Varma, P., Sebastian, P., Gao, R., Noto, T., Lara, A. H., Wallis, J. D., Knight, R. T., Shestyuk, A., & Voytek, B. (2020). Parameterizing neural power spectra into periodic and aperiodic components. Nature Neuroscience, 23(12), 1655–1665. 10.1038/s41593-020-00744-x

Dugué, L., & Chavane, F. (2025). Traveling waves across scales: Different mechanisms but same canonical computation? ELife, 14. 10.7554/eLife.106753

Ebersole, J. S. (1997). Defining epileptogenic foci: past, present, future. Journal of Clinical Neurophysiology, 14(6), 470–483. 10.1097/00004691-199711000-00003

Ebersole, J. S. (2000). Noninvasive localization of epileptogenic foci by EEG source modeling. Epilepsia, 41 *Suppl 3*, S24–33. 10.1111/j.1528-1157.2000.tb01531.x

Elul, R. (1971). The genesis of the EEG. International Review of Neurobiology, 15, 227–272. 10.1016/S0074-7742(08)60333-5

Evans, A. C., Collins, D. L., Mills, S. R., Brown, E. D., Kelly, R. L., & Peters, T. M. (1993). 3D statistical neuroanatomical models from 305 MRI volumes. 1993 IEEE Conference Record Nuclear Science Symposium and Medical Imaging Conference, 1813–1817. 10.1109/NSSMIC.1993.373602

Fonov, V. S., Evans, A. C., Botteron, K., Almli, C. R., McKinstry, R. C., Collins, D. L., & Brain Development Cooperative Group. (2011). Unbiased average age-appropriate atlases for pediatric studies. Neuroimage, 54(1), 313–327. 10.1016/j.neuroimage.2010.07.033

Friston, K. J. (1998). Modes or models: a critique on independent component analysis for fMRI. Trends in Cognitive Sciences, 2(10), 373–375. 10.1016/s1364-6613(98)01227-3

Fuchs, M., Kastner, J., Tech, R., Wagner, M., & Gasca, F. (2017). MEG and EEG dipole clusters from extended cortical sources. Biomedical Engineering Letters, 7(3), 185–191. 10.1007/s13534-017-0019-2

Garcia-Marin, V., Kelly, J. G., & Hawken, M. J. (2019). Major feedforward thalamic input into layer 4C of primary visual cortex in primate. Cerebral Cortex, 29(1), 134–149. 10.1093/cercor/bhx311

Getzmann, S., Gajewski, P. D., Schneider, D., & Wascher, E. (2024). Resting-state EEG data before and after cognitive activity across the adult lifespan and a 5-year follow-up. Scientific Data, 11(1), 988. 10.1038/s41597-024-03797-w

Halgren, M., Ulbert, I., Bastuji, H., Fabó, D., Erőss, L., Rey, M., Devinsky, O., Doyle, W. K., Mak-McCully, R., Halgren, E., Wittner, L., Chauvel, P., Heit, G., Eskandar, E., Mandell, A., & Cash, S. S. (2019). The generation and propagation of the human alpha rhythm. Proceedings of the National Academy of Sciences of the United States of America, 116(47), 23772–23782. 10.1073/pnas.1913092116

Halnes, G., Ness, T. V., Næss, S., Hagen, E., Pettersen, K. H., & Einevoll, G. T. (2024). Electric brain signals: foundations and applications of biophysical modeling. Cambridge University Press. 10.1017/9781009039826

Hari, R., Salmelin, R., Mäkelä, J. P., Salenius, S., & Helle, M. (1997). Magnetoencephalographic cortical rhythms. International Journal of Psychophysiology, 26(1–3), 51–62. 10.1016/S0167-8760(97)00755-1

Hari, R. (1990). The neuromagnetic method in the study of the human auditory cortex. Advances in Audiology, 6, 222–282.

Hashiguchi, K., Morioka, T., Yoshida, F., Miyagi, Y., Nagata, S., Sakata, A., & Sasaki, T. (2007). Correlation between scalp-recorded electroencephalographic and electrocorticographic activities during ictal period. Seizure, 16(3), 238–247. 10.1016/j.seizure.2006.12.010

Huang, J.-Y., Bock, O., Naveilhan, C., Ramanoël, S., Faßbender, R. V., Memmert, D., & Onur, O. A. (2026). Cortical dynamics are differentially associated with decision-making strategies in human wayfinding: An EEG study. Cognitive, Affective & Behavioral Neuroscience, 26(1), 120–138. 10.3758/s13415-025-01357-2

Hughes, J. R. (1995). The phenomenon of travelling waves: a review. Clinical EEG (Electroencephalography), 26(1), 1–6.

Hyvärinen, A., Karhunen, J., & Oja, E. (2001). Independent Component Analysis (1st ed., p. 504). Wiley-Interscience.

Ikeda, A., Taki, W., Kunieda, T., Terada, K., Mikuni, N., Nagamine, T., Yazawa, S., Ohara, S., Hori, T., Kaji, R., Kimura, J., & Shibasaki, H. (1999). Focal ictal direct current shifts in human epilepsy as studied by subdural and scalp recording. Brain: A Journal of Neurology, 122 ( *Pt 5*), 827–838. 10.1093/brain/122.5.827

Jeong, S.-O., Ko, T.-W., & Moon, H.-T. (2002). Time-delayed spatial patterns in a two-dimensional array of coupled oscillators. Physical Review Letters, 89(15), 154104. 10.1103/PhysRevLett.89.154104

Jungnickel, E., & Gramann, K. (2016). Mobile Brain/Body Imaging (MoBI) of Physical Interaction with Dynamically Moving Objects. Frontiers in Human Neuroscience, 10, 306. 10.3389/fnhum.2016.00306

Jung, T.-P., Makeig, S., Humphries, C., Lee, T. W., McKeown, M. J., Iragui, V., & Sejnowski, T. J. (2000). Removing electroencephalographic artifacts by blind source separation. Psychophysiology, 37(2), 163–178. 10.1111/1469-8986.3720163

Jung, T.-P., Makeig, S., McKeown, M. J., Bell, A. J., Lee, T.-W., & Sejnowski, T. J. (2001). Imaging Brain Dynamics Using Independent Component Analysis. Proceedings of the IEEE. Institute of Electrical and Electronics Engineers, 89(7), 1107–1122. 10.1109/5.939827

Jung, T.-P., Makeig, S., Westerfield, M., Townsend, J., Courchesne, E., & Sejnowski, T. J. (2000). Removal of eye activity artifacts from visual event-related potentials in normal and clinical subjects. Clinical Neurophysiology, 111(10), 1745–1758. 10.1016/S1388-2457(00)00386-2

Jung, T.-P., Makeig, S., Westerfield, M., Townsend, J., Courchesne, E., & Sejnowski, T. J. (2001). Analysis and visualization of single-trial event-related potentials. Human Brain Mapping, 14(3), 166–185. 10.1002/hbm.1050

Kanari, L., Ramaswamy, S., Shi, Y., Morand, S., Meystre, J., Perin, R., Abdellah, M., Wang, Y., Hess, K., & Markram, H. (2019). Objective morphological classification of neocortical pyramidal cells. Cerebral Cortex, 29(4), 1719–1735. 10.1093/cercor/bhy339

Kim, H., Chang, C.-Y., Kothe, C., Iversen, J. R., & Miyakoshi, M. (2025). Juggler’s ASR: Unpacking the principles of artifact subspace reconstruction for revision toward extreme MoBI. Journal of Neuroscience Methods, 420, 110465. 10.1016/j.jneumeth.2025.110465

Kim, H., Luo, J., Chu, S., Cannard, C., Hoffmann, S., & Miyakoshi, M. (2023). ICA’s bug: How ghost ICs emerge from effective rank deficiency caused by EEG electrode interpolation and incorrect re-referencing. Frontiers in Signal Processing, 3. 10.3389/frsip.2023.1064138

Kobayashi, K., Yoshinaga, H., Ohtsuka, Y., & Gotman, J. (2005). Dipole modeling of epileptic spikes can be accurate or misleading. Epilepsia, 46(3), 397–408. 10.1111/j.0013-9580.2005.31404.x

Kothe, C., & Jung, T.-P. (2016). Artifact removal techniques with signal reconstruction (Patent No. US20160113587A1).

Kothe, C., & Makeig, S. (2013). BCILAB: a platform for brain-computer interface development. Journal of Neural Engineering, 10(5), 056014. 10.1088/1741-2560/10/5/056014

Lahijanian, M., Aghajan, H., & Vahabi, Z. (2024). Auditory gamma-band entrainment enhances default mode network connectivity in dementia patients. Scientific Reports, 14(1), 13153. 10.1038/s41598-024-63727-z

Latawiec, D., Martin, K. A., & Meskenaite, V. (2000). Termination of the geniculocortical projection in the striate cortex of macaque monkey: a quantitative immunoelectron microscopic study. The Journal of Comparative Neurology, 419(3), 306–319. 10.1002/(sici)1096-9861(20000410)419:3<306::aid-cne4>3.0.co;2-2

Lin, C.-T., Chiu, T.-C., Wang, Y.-K., Chuang, C.-H., & Gramann, K. (2018). Granger causal connectivity dissociates navigation networks that subserve allocentric and egocentric path integration. Brain Research, 1679, 91–100. 10.1016/j.brainres.2017.11.016

Makeig, S., Bell, A., Jung, T.-P., & Sejnowski, T. (1996). Independent component analysis of electroencephalographic data. Advances in Neural Information Processing Systems 8, 145–151.

Makeig, S., Debener, S., Onton, J., & Delorme, A. (2004). Mining event-related brain dynamics. Trends in Cognitive Sciences, 8(5), 204–210. 10.1016/j.tics.2004.03.008

Makeig, S., Delorme, A., Westerfield, M., Jung, T.-P., Townsend, J., Courchesne, E., & Sejnowski, T. J. (2004). Electroencephalographic brain dynamics following manually responded visual targets. PLoS Biology, 2(6), e176. 10.1371/journal.pbio.0020176

Makeig, S., Jung, T.-P., Bell, A. J., Ghahremani, D., & Sejnowski, T. J. (1997). Blind separation of auditory event-related brain responses into independent components. Proceedings of the National Academy of Sciences of the United States of America, 94(20), 10979–10984. 10.1073/pnas.94.20.10979

Makeig, S., & Onton, J. (2011). ERP features and EEG dynamics. Oxford University Press. 10.1093/oxfordhb/9780195374148.013.0035

Makeig, S., Westerfield, M., Jung, T.-P., Covington, J., Townsend, J., Sejnowski, T. J., & Courchesne, E. (1999). Functionally independent components of the late positive event-related potential during visual spatial attention. The Journal of Neuroscience, 19(7), 2665–2680. 10.1523/JNEUROSCI.19-07-02665.1999

Makeig, S., Westerfield, M., Jung, T.-P., Enghoff, S., Townsend, J., Courchesne, E., & Sejnowski, T. J. (2002). Dynamic brain sources of visual evoked responses. Science, 295(5555), 690–694. 10.1126/science.1066168

Makeig, S., Westerfield, M., Townsend, J., Jung, T.-P., Courchesne, E., & Sejnowski, T. J. (1999). Functionally independent components of early event-related potentials in a visual spatial attention task. Philosophical Transactions of the Royal Society of London. Series B, Biological Sciences, 354(1387), 1135–1144. 10.1098/rstb.1999.0469

Mazziotta, J. C., Toga, A. W., Evans, A., Fox, P., & Lancaster, J. (1995). A probabilistic atlas of the human brain: theory and rationale for its development. The International Consortium for Brain Mapping (ICBM). Neuroimage, 2(2), 89–101. 10.1006/nimg.1995.1012

Miyakoshi, M., Kim, H., De Stefano, L. A., Schmitt, L. M., Norris, J. E., Ethridge, L. E., Erickson, C. A., & Pedapati, E. V. (2025). Hyper-extralemniscal model of Fragile X syndrome. Cerebral Cortex, 35(6). 10.1093/cercor/bhaf141

Miyakoshi, M., Kim, H., Nakanishi, M., Palmer, J., & Kanayama, N. (2023). One out of ten independent components shows flipped polarity with poorer data quality: EEG database study. Human Brain Mapping. 10.1002/hbm.26540

Miyakoshi, M. (2023). Artifact subspace reconstruction (ASR): A candidate for a dream solution for EEG studies, sleep or awake. Sleep, zsad241. 10.1093/sleep/zsad241

Mognon, A., Jovicich, J., Bruzzone, L., & Buiatti, M. (2011). ADJUST: An automatic EEG artifact detector based on the joint use of spatial and temporal features. Psychophysiology, 48(2), 229–240. 10.1111/j.1469-8986.2010.01061.x

Mouraux, A., & Iannetti, G. D. (2009). Nociceptive laser-evoked brain potentials do not reflect nociceptive-specific neural activity. Journal of Neurophysiology, 101(6), 3258– 3269. 10.1152/jn.91181.2008

Muller, L., Chavane, F., Reynolds, J., & Sejnowski, T. J. (2018). Cortical travelling waves: mechanisms and computational principles. Nature Reviews. Neuroscience, 19(5), 255–268. 10.1038/nrn.2018.20

Næss, S., Chintaluri, C., Ness, T. V., Dale, A. M., Einevoll, G. T., & Wójcik, D. K. (2017). Corrected Four-Sphere Head Model for EEG Signals. Frontiers in Human Neuroscience, 11, 490. 10.3389/fnhum.2017.00490

Nolan, H., Whelan, R., & Reilly, R. B. (2010). FASTER: Fully Automated Statistical Thresholding for EEG artifact Rejection. Journal of Neuroscience Methods, 192(1), 152–162. 10.1016/j.jneumeth.2010.07.015

Nunez, P. L., & Srinivasan, R. (2006). Electric fields of the brain. Oxford University Press, Inc. 10.1093/acprof:oso/9780195050387.001.0001

O’Kusky, J., & Colonnier, M. (1982). A laminar analysis of the number of neurons, glia, and synapses in the adult cortex (area 17) of adult macaque monkeys. The Journal of Comparative Neurology, 210(3), 278–290. 10.1002/cne.902100307

Onton, J., Delorme, A., & Makeig, S. (2005). Frontal midline EEG dynamics during working memory. Neuroimage, 27(2), 341–356. 10.1016/j.neuroimage.2005.04.014

Onton, J., & Makeig, S. (2006). Information-based modeling of event-related brain dynamics. Progress in Brain Research, 159, 99–120. 10.1016/S0079-6123(06)59007-7

Onton, J., Westerfield, M., Townsend, J., & Makeig, S. (2006). Imaging human EEG dynamics using independent component analysis. Neuroscience and Biobehavioral Reviews, 30(6), 808–822. 10.1016/j.neubiorev.2006.06.007

Oostenveld, R., Fries, P., Maris, E., & Schoffelen, J.-M. (2011). FieldTrip: Open source software for advanced analysis of MEG, EEG, and invasive electrophysiological data. Computational Intelligence and Neuroscience, 2011, 156869. 10.1155/2011/156869

Oostenveld, R., & Praamstra, P. (2001). The five percent electrode system for high-resolution EEG and ERP measurements. Clinical Neurophysiology, 112(4), 713–719. 10.1016/S1388-2457(00)00527-7

Pakkenberg, B., & Gundersen, H. J. (1997). Neocortical neuron number in humans: effect of sex and age. The Journal of Comparative Neurology, 384(2), 312–320. 10.1002/(SICI)1096-9861(19970728)384:2<312::AID-CNE10>3.0.CO;2-K

Palmer, J., Kreutz-delgado, K., & Makeig, S. (2016). AMICA: An Adaptive Mixture of Independent Component Analyzers with Shared Components.

Peters, A., & Payne, B. R. (1993). Numerical relationships between geniculocortical afferents and pyramidal cell modules in cat primary visual cortex. Cerebral Cortex, 3(1), 69–78. 10.1093/cercor/3.1.69

Phillips, C., Kline, J., Stanley, C. J., Bulea, T. C., & Damiano, D. L. (2023). Children with bilateral cerebral palsy exhibit bimanual asymmetric motor deficits and EEG evidence of dominant sensorimotor hemisphere overreliance during reaching. Neurorehabilitation and Neural Repair, 37(9), 617–627. 10.1177/15459683231195044

Piazza, C., Miyakoshi, M., Akalin Acar, Z., Cantiani, C., Reni, G., Bianchi, A. M., & Makeig, S. (2016). An Automated Function for Identifying EEG Independent Components Representing Bilateral Source Activity. In E. Kyriacou, S. Christofides, & C. S. Pattichis (Eds.), XIV Mediterranean Conference on Medical and Biological Engineering and Computing 2016 (Vol. 57, pp. 105–109). Springer International Publishing. 10.1007/978-3-319-32703-7_22

Pion-Tonachini, L., Kreutz-Delgado, K., & Makeig, S. (2019). ICLabel: An automated electroencephalographic independent component classifier, dataset, and website. Neuroimage, 198, 181–197. 10.1016/j.neuroimage.2019.05.026

Scherg, M., Berg, P., Nakasato, N., & Beniczky, S. (2019). Taking the EEG back into the brain: the power of multiple discrete sources. Frontiers in Neurology, 10, 855. 10.3389/fneur.2019.00855

Schoonover, C. E., Tapia, J.-C., Schilling, V. C., Wimmer, V., Blazeski, R., Zhang, W., Mason, C. A., & Bruno, R. M. (2014). Comparative strength and dendritic organization of thalamocortical and corticocortical synapses onto excitatory layer 4 neurons. The Journal of Neuroscience, 34(20), 6746–6758. 10.1523/JNEUROSCI.0305-14.2014

Somervail, R., Perovic, S., Bufacchi, R. J., Caminiti, R., & Iannetti, G. D. (2026). A two-system theory of sensory-evoked brain responses. Brain: A Journal of Neurology, 149(5), 1438–1451. 10.1093/brain/awaf402

Stark, A. K., Toft, M. H., Pakkenberg, H., Fabricius, K., Eriksen, N., Pelvig, D. P., Møller, M., & Pakkenberg, B. (2007). The effect of age and gender on the volume and size distribution of neocortical neurons. Neuroscience, 150(1), 121–130. 10.1016/j.neuroscience.2007.06.062

Stepanyants, A., Martinez, L. M., Ferecskó, A. S., & Kisvárday, Z. F. (2009). The fractions of short- and long-range connections in the visual cortex. Proceedings of the National Academy of Sciences of the United States of America, 106(9), 3555–3560. 10.1073/pnas.0810390106

Stettler, D. D., Das, A., Bennett, J., & Gilbert, C. D. (2002). Lateral connectivity and contextual interactions in macaque primary visual cortex. Neuron, 36(4), 739–750. 10.1016/s0896-6273(02)01029-2

Symeonidou, E.-R., & Ferris, D. P. (2023). Visual occlusions result in phase synchrony within multiple brain regions involved in sensory processing and balance control. IEEE Transactions on Neural Systems and Rehabilitation Engineering, 31, 3772– 3780. 10.1109/TNSRE.2023.3317055

Tao, J. X., Baldwin, M., Ray, A., Hawes-Ebersole, S., & Ebersole, J. S. (2007). The impact of cerebral source area and synchrony on recording scalp electroencephalography ictal patterns. Epilepsia, 48(11), 2167–2176. 10.1111/j.1528-1167.2007.01224.x

Tao, J. X., Ray, A., Hawes-Ebersole, S., & Ebersole, J. S. (2005). Intracranial EEG substrates of scalp EEG interictal spikes. Epilepsia, 46(5), 669–676. 10.1111/j.1528-1167.2005.11404.x

Tsai, A. C., Jung, T.-P., Chien, V. S. C., Savostyanov, A. N., & Makeig, S. (2014). Cortical surface alignment in multi-subject spatiotemporal independent EEG source imaging. Neuroimage, 87, 297–310. 10.1016/j.neuroimage.2013.09.045

Tzourio-Mazoyer, N., Landeau, B., Papathanassiou, D., Crivello, F., Etard, O., Delcroix, N., Mazoyer, B., & Joliot, M. (2002). Automated anatomical labeling of activations in SPM using a macroscopic anatomical parcellation of the MNI MRI single-subject brain. Neuroimage, 15(1), 273–289. 10.1006/nimg.2001.0978

Udeigwe, L. C., & Ermentrout, G. B. (2015). Waves and patterns on regular graphs. SIAM Journal on Applied Dynamical Systems, 14(2), 1102–1129. 10.1137/140969488

Van Essen, D. C., & Drury, H. A. (1997). Structural and functional analyses of human cerebral cortex using a surface-based atlas. The Journal of Neuroscience, 17(18), 7079–7102. 10.1523/JNEUROSCI.17-18-07079.1997

Walter, W. G., Cooper, R., Aldridge, V. J., Mccallum, W. C., & Winter, A. L. (1964). Contingent negative variation: an electric sign of sensorimotor association and expectancy in the human brain. Nature, 203, 380–384.

Walter, W. G. (1964). The convergence and interaction of visual, auditory, and tactile responses in human nonspecific cortex. Annals of the New York Academy of Sciences, 112, 320–361. 10.1111/j.1749-6632.1964.tb26760.x

Whitmer, D., Worrell, G., Stead, M., Lee, I. K., & Makeig, S. (2010). Utility of independent component analysis for interpretation of intracranial EEG. Frontiers in Human Neuroscience, 4, 184. 10.3389/fnhum.2010.00184

Winkler, I., Haufe, S., & Tangermann, M. (2011). Automatic classification of artifactual ICA-components for artifact removal in EEG signals. Behavioral and Brain Functions, 7, 30. 10.1186/1744-9081-7-30

Zilles, K., Armstrong, E., Schleicher, A., & Kretschmann, H. J. (1988). The human pattern of gyrification in the cerebral cortex. Anatomy and Embryology, 179(2), 173–179. 10.1007/BF00304699

