## Supplement_mm for "Small or large patches? History, biophysics, and an empirical test of a core assumption in EEG independent component analysis"

### 1. Alignment between the AAL template head model and the EEGLAB default head model.

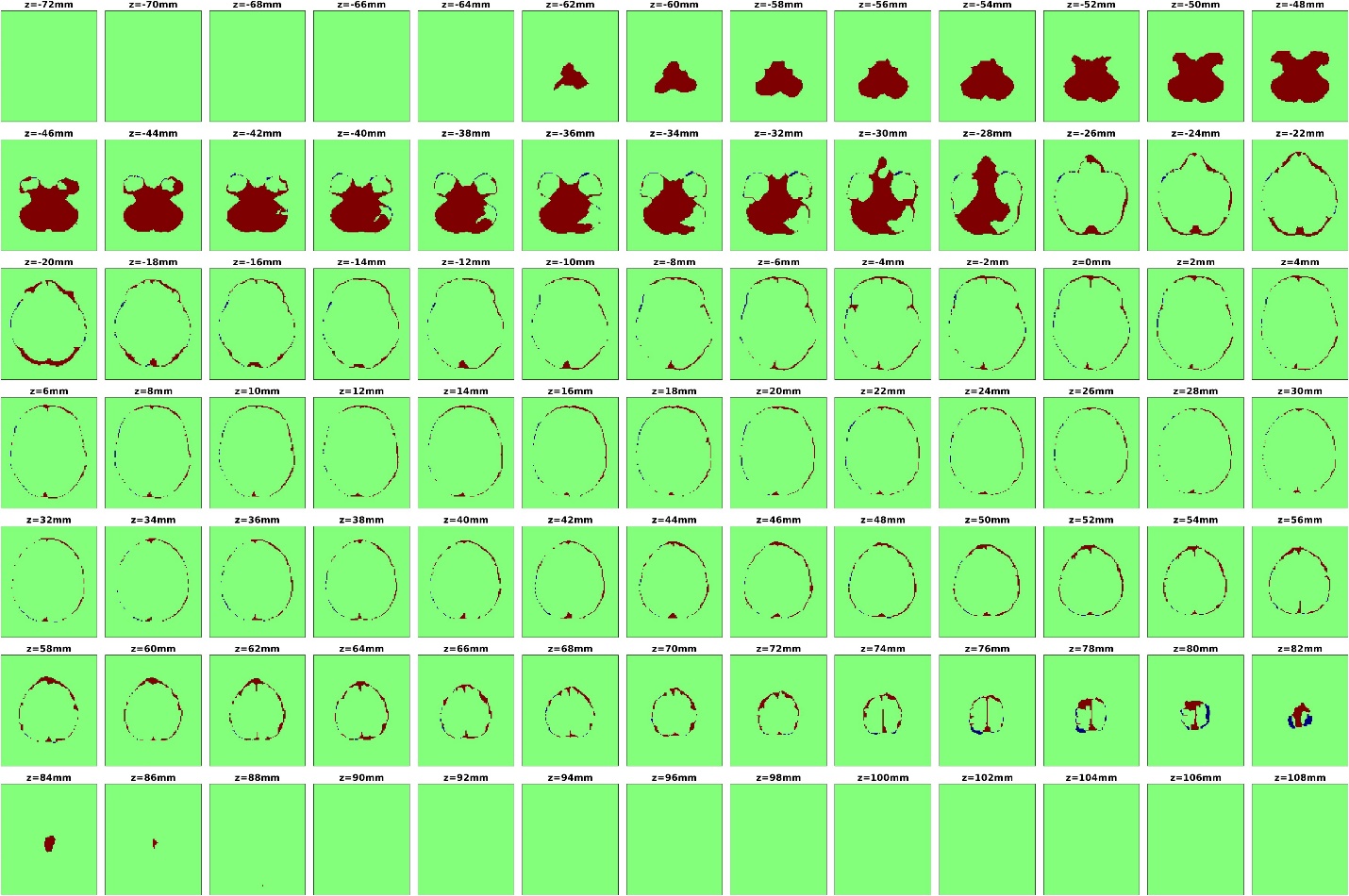

**Figure S1. Alignment between the AAL template head model and the EEGLAB default head model.** To enable anatomical labeling of EEGLAB-derived dipole fits using the AAL atlas, the AAL head model was spatially expanded to match the scale of the EEGLAB default head model. The original minor mismatch in size and shape is illustrated across 91 axial slices. The transformed (expanded) AAL volume preserves anatomical topology while achieving voxel-wise correspondence with the EEGLAB head model. This alignment step allows dipole fitting results to be mapped to AAL anatomical labels in a consistent stereotaxic space. Red, EEGLAB > AAL; Blue, AAL > EEGLAB.

### 2. Residual variance as a function of UDL radius after dipole fitting across different BSCR values.

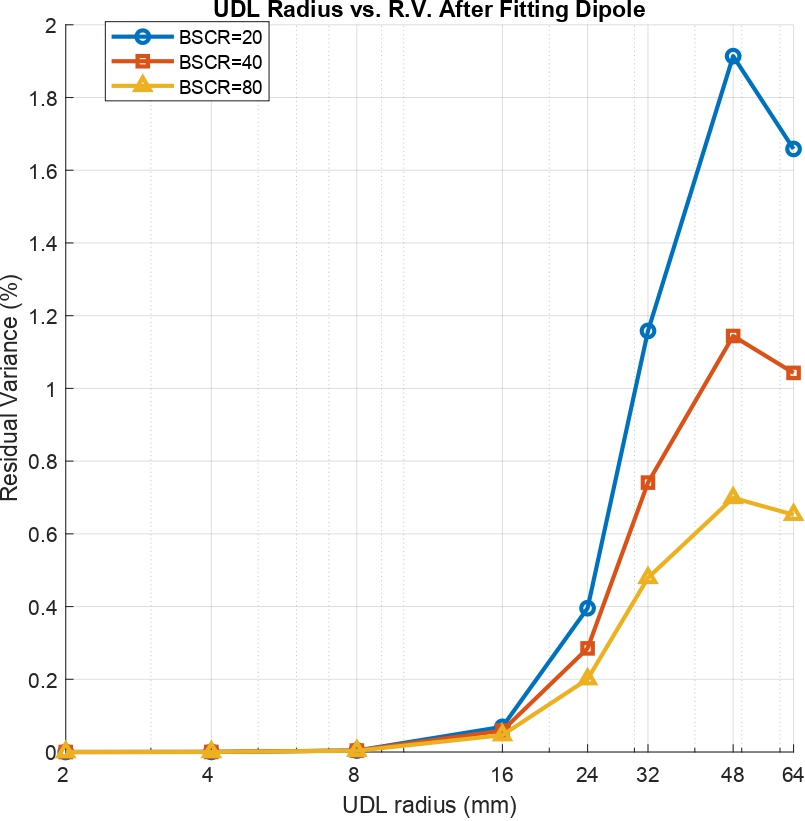

**Figure S2. Residual variance as a function of UDL radius after dipole fitting across different BSCR values.** The plot shows residual variance (%) versus UDL radius (mm) for three BSCR conditions (20, 40, and 80). All curves remain near zero for small UDL radii (2–16 mm), indicating excellent single-dipole fits. Beyond approximately 24 mm, residual variance increases with UDL radius, with the largest increase observed at BSCR = 20 (blue circles, reaching approximately 1.9% at 48 mm), followed by BSCR = 40 (orange squares) and BSCR = 80 (yellow triangles, reaching approximately 0.7% at 48 mm). Residual variance decreases slightly at 64 mm across all three conditions. These results indicate that increasing the spatial extent of a distributed source reduces the goodness of fit of a single equivalent dipole, particularly at lower BSCR values. Nevertheless, residual variance remains below 2% across all conditions, demonstrating that even spatially extended generators can produce scalp potential distributions that are well approximated by a single dipole.

**EEG_Clustering Plugin: Implementation and Demonstration**

**Overview**

For comprehensive source-level data preparation, clustering, and analysis, we developed a custom graphical user interface (GUI)-based EEGLAB plugin named 'EEG_Clustering' by transforming a MATLAB Designer application (EEG_Clustering.mlapp) into the EEGLAB plugin framework. The plugin accommodates 1 to 3 groups and 2 to 20 clusters, offering automated workflows for extracting detailed dipole information and constructing group-separated and combined cluster-specific datasets for further analysis. The implementation supports the visualization of source-level metrics, including Spectra, Event-Related Spectral Perturbations (ERSP), Inter-Trial Coherence (ITC), dipole, and dipole density plots, alongside the export of numerical data for cluster-specific statistical evaluation. The example presented here is based on a simulated dataset consisting of two groups (HS and PS) with a total of 33 participants.

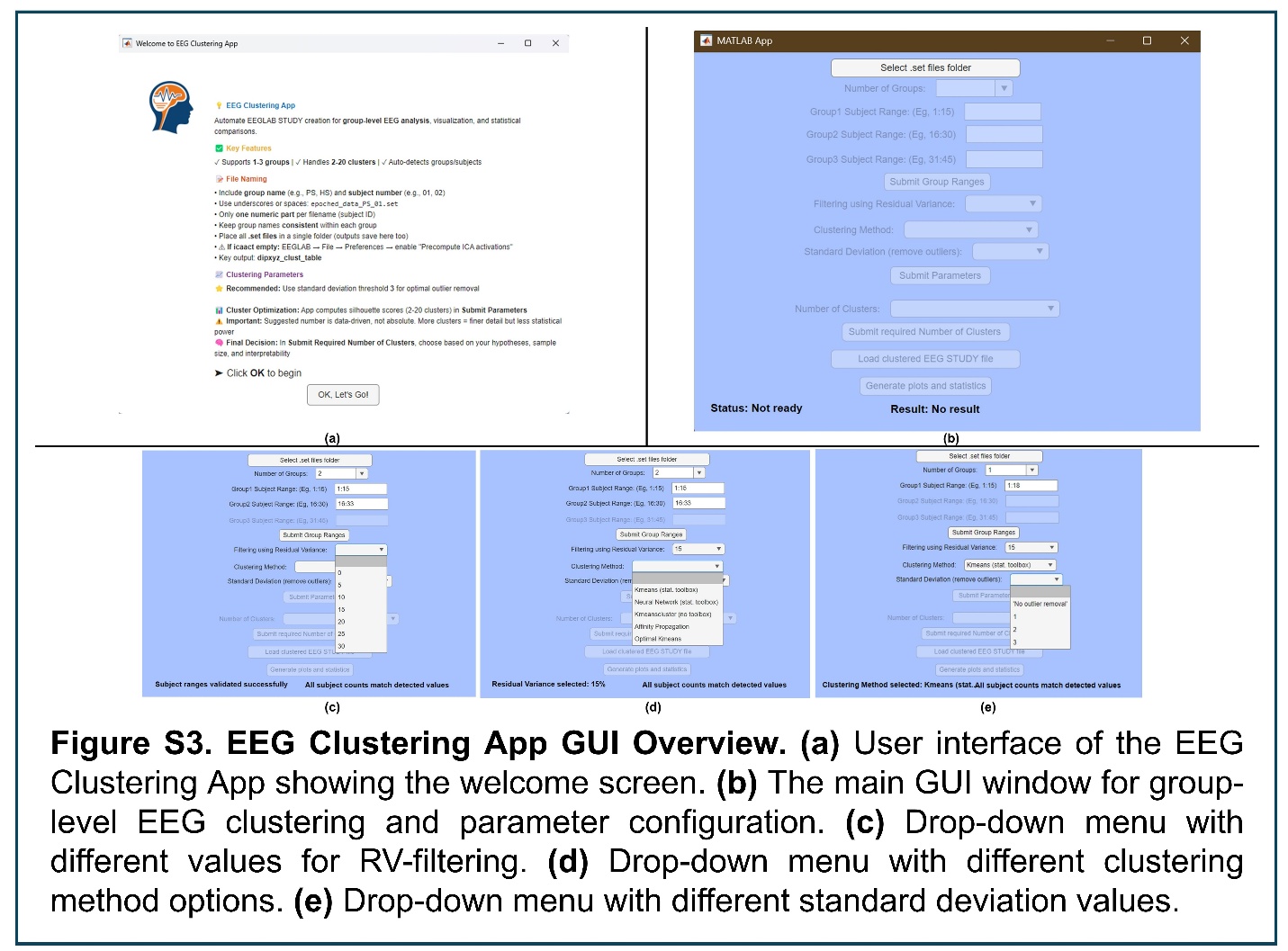

### 3. EEG Clustering Application GUI Overview.

**Welcome Screen and GUI Initialization**

Upon launching the EEG_Clustering application, a startup function triggers the display of a modal welcome screen (**Fig. S3a**) designed to improve user interaction. This screen contains instructional text outlining the plugin's capabilities, including support for 1 to 3 groups and 2 to 20 clusters, and detailed file-naming conventions using delimiters such as underscores. A confirmation button labeled "OK, Let's Go!" allows users to close the welcome screen and proceed to the main GUI. The main interface adopts an EEGLAB-based design with a light blue background and standardized control styling (**Fig. S3a**). During initialization, the startup function also configures essential parameters, such as file selection mechanisms and status labels, ensuring the application is ready for subsequent EEG data processing and clustering analysis.

**Study File Selection and Dataset Loading**

Upon executing the "Select Study Files" function, the application prompts the user to select a directory containing epoched .set files. The selected folder is validated, and its contents are scanned for EEG datasets. Files not conforming to the expected naming convention (e.g., missing group prefix or subject number) trigger descriptive warning messages. The parsing algorithm identifies 1–3 experimental groups, extracts subject IDs, and organizes the results into a structured table sorted by group and subject number. For the simulated dataset, the selection of a folder containing .set files was successfully executed through a directory interface. Filename parsing accurately identified two experimental groups (HS and PS) across a total of 33 subjects, with 15 subjects in the HS group and 18 subjects in the PS group. All files were valid, and no warnings were generated during parsing.

EEGLAB is initialized automatically, and each dataset is sequentially loaded. If any dataset lacks ICA activation data (icaact), it is recomputed from the ICA weights and sphere matrices to ensure completeness. In the test example, EEGLAB initialized automatically and successfully loaded all datasets. ICA activation data were present for 30 datasets and recomputed for 3 datasets where it was missing. Following successful loading and validation of all datasets, a non-clustered EEGLAB STUDY is created using pop_study. The STUDY is populated with consistent subject and group metadata and undergoes component-level precomputation using std_precomp. Finally, the STUDY is saved to the selected directory, and the application enables the "Number of Groups" dropdown within the main graphical user interface (GUI) (**Fig. S3b**), ensuring a smooth transition from dataset selection to an initialized analysis environment.

**Group Selection and Range Specification**

The "Number of Groups" section within the EEG_Clustering interface enables users to specify the number of experimental groups (1, 2, or 3) via a dropdown menu. The selected value is internally parsed and validated to ensure compliance with the supported range. Invalid inputs, such as values outside the 1–3 range, immediately trigger an error message, reset the dropdown to the default value (1), and disable further progression. Upon valid selection, the application compares the user-specified group count against the group count automatically detected during file selection. In the test example, the user selected "2" to match the app-detected structure, which had previously identified two experimental groups: HS and PS. The system confirmed this match and updated the status and result indicators accordingly. The graphical interface dynamically enabled the appropriate range input fields for two groups, while keeping the third field inactive, guiding the user toward consistent input (**Fig. S3b**).

In the "Group Ranges" section, users define subject ID ranges corresponding to each group selected. Upon pressing the "Submit" button, the application initiates a validation process. First, it checks that the user-entered ranges are syntactically correct (e.g., "1:15"), properly formatted as numeric arrays, and do not contain multiple or ambiguous inputs. Second, the system ensures that the numerical values fall within the detected subject ID bounds established during dataset parsing. Ranges that are incomplete, incorrectly formatted, or exceed the available subject IDs trigger descriptive error messages and immediately terminate execution. For the test dataset, subject ID ranges were entered as "1:15" for Group 1 (HS) and "16:33" for Group 2 (PS), matching the actual subject distribution parsed from the dataset. The application validated the format, converted the ranges into numerical arrays, and confirmed that the subject counts per group (15 and 18, respectively) were consistent with the detected values. Upon successful validation, a status message indicated readiness for the next configuration step, and the interface activated the dropdowns for residual variance filtering, clustering method selection, and standard deviation for outlier removal (**Fig. S3b**).

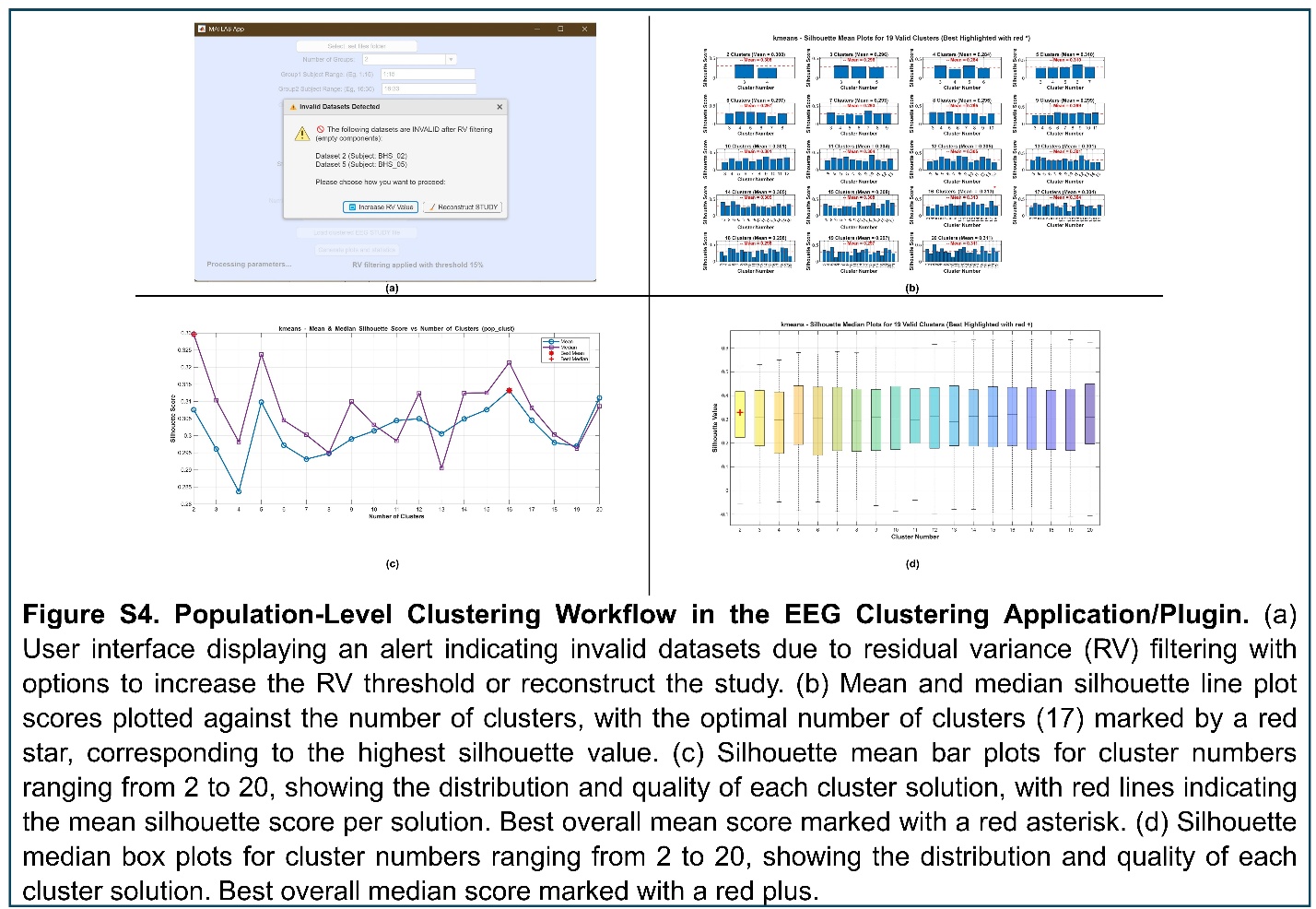

### 4. Population-Level Clustering Workflow in the EEG Clustering Application/Plugin.

**Clustering Parameter Configuration**

The EEG_Clustering plugin enables users to filter EEG data by selecting a residual variance (RV) threshold via a dropdown menu in the graphical user interface (**Fig. S3c**). For example, choosing 15% converts the input to a decimal (0.15) and stores it in the app.residualVariance variable. The plugin validates that the input is numeric and between 0 and 100%. If invalid (e.g., 150% or non-numeric), an alert dialog notifies the user, the status label displays "Error: Invalid residual variance," the result label shows "Please select a value between 0 and 100%," and processing halts. For valid inputs, the status label updates (e.g., "Residual Variance selected: 15%"), and the selection is logged to the command window for user confirmation. In the test example, when users selected a residual variance threshold of 15% via the GUI (**Fig. S3c**), the plugin stored it as app.residualVariance = 0.15 and updated the status label to "Residual Variance selected: 15%."

Users select a clustering method via a dropdown menu (**Fig. S3d**) from options: "Kmeans (stat. toolbox)," "Neural Network (stat. toolbox)," "Kmeanscluster (no toolbox)," "Affinity Propagation," and "Optimal Kmeans." The selection is stored in app.clusteringMethod, logged, and updates the status label (e.g., "Clustering Method selected: Kmeans (stat. toolbox)"). For "Kmeans (stat. toolbox)," "Kmeanscluster (no toolbox)," and "Optimal Kmeans," the standard deviation dropdown is enabled, but the "Submit Parameters" button remains disabled until a threshold is selected. For "Neural Network (stat. toolbox)" or "Affinity Propagation," a warning dialog indicates no outlier removal support, offering to continue (disabling the standard deviation dropdown, enabling "Submit Parameters") or switch algorithms (resetting dropdowns to defaults, keeping "Submit Parameters" disabled). The plugin logs the choice, providing clear feedback without additional checks, allowing users to move to the next step. In the test example, selecting "Kmeans (stat. toolbox)" via the GUI (**Fig. S3d**) set app.clusteringMethod accordingly, with the status label updating to "Clustering Method selected: Kmeans (stat. toolbox)."

Users set a threshold for outlier removal via a dropdown menu (**Fig. S3e**). For example, selecting a value of 3 stores it as app.stdDevThreshold. Non-numeric inputs default to 0 (no outlier removal). The plugin logs the threshold to the command window and updates the status label (e.g., "Selected Standard Deviation Threshold: 3" or "No outlier removal"). This action enables the "Submit Parameters" button in the GUI, allowing users to proceed to clustering without further validation. In the test example, choosing an outlier threshold of 3 via the GUI (**Fig. S3e**) set app.stdDevThreshold to 3 and updated the status label to "Standard Deviation Threshold selected: 3." The "Submit Parameters" button was enabled, and the selection was logged, streamlining clustering preparation.

**Clustering Execution and Study Reconstruction**

Upon clicking the "Submit Parameters" button (**Fig. S3b**), the plugin initiates clustering by logging group prefixes (e.g., HS, PS) and subject ranges to the command window, setting the status label to "Processing parameters...". It verifies that the STUDY and ALLEEG data structures are loaded. If either is missing, an alert dialog displays "STUDY or ALLEEG data is not loaded. Please load a STUDY file first," the status label updates to "Error: Data not loaded," the result label shows "Load a STUDY file first," and processing halts. Valid parameters (e.g., RV = 0.15, clustering method = Kmeans, threshold = 3) are checked, with the clustering method mapped to an EEGLAB-compatible algorithm (e.g., "Kmeans (stat. toolbox)" to kmeans). The plugin counts datasets (e.g., 33 datasets: 15 HS, 18 PS) and dipoles (e.g., 601) using multiple methods to ensure consistency. Residual variance filtering is applied using the selected threshold, updating the result label (e.g., "RV filtering applied with threshold 15%"). For the simulated dataset (33 datasets: 15 HS, 18 PS; 601 dipoles), the plugin verified that STUDY and ALLEEG were loaded and confirmed 33 datasets and 601 dipoles before RV filtering, applied RV filtering, and updated the result label to "RV filtering applied with threshold 15%."

After RV filtering, the plugin assesses data integrity by recounting datasets and dipoles, logging results to the command window. For example, a 15% RV threshold may yield 31 valid datasets (from 33) and 335 dipoles (from 365 after RV filtering). If mismatches occur (e.g., 31 valid vs. 33 total datasets, with HS_02 and HS_05 invalid), a modal dialog (**Fig. S4a**) offers two options: (1) Increase RV Value: A confirmation dialog prompts the user (e.g., "If you click Yes, the app will restart and then select RV > 15%... Proceed?"). If confirmed, the status label shows "Resetting...", UI elements are disabled, the result label updates to "Please start by selecting a folder to proceed," and an alert guides folder reselection. If cancelled, the status label shows "Action Cancelled." (2) Reconstruct STUDY: The plugin excludes invalid datasets (e.g., HS_02, HS_05), reloads valid datasets (e.g., 31) into ALLEEG, recreates the STUDY, and precomputes components, yielding 335 dipoles. The result label updates to "STUDY reconstructed successfully with 31 datasets." Mismatches (e.g., dataset vs. dipole counts) halt processing, with the result label indicating the issue (e.g., "Dataset and dipole counts do not match"). In the test example, after applying a 15% RV threshold, the plugin counted 31 valid datasets (HS_02 and HS_05 invalid) and components (335 and 365 comps), logging details to the command window. The mismatch (31 vs. 33 datasets; and 335 vs. 365 components) triggered the dialog (**Fig. S4a**). Selecting "Reconstruct STUDY" excluded HS_02 and HS_05, reloaded 31 datasets, and recreated the STUDY, yielding 335 dipoles (30 from invalid datasets removed), with the result label updating to "STUDY reconstructed successfully with 31 datasets."

Pre-clustering is performed using std_preclust with dipole normalization and weighting, logging "Performing pre-clustering..." to the command window. The plugin validates component counts and dipole locations, ensuring no anomalies (e.g., NaN values). Post-pre-clustering, it recounts datasets (e.g., 31) and dipoles (e.g., 335). If mismatches occur (e.g., 31 vs. 33 datasets), invalid datasets are logged (e.g., "Dataset 2 (Subject: HS_02)"), and the **Fig. S4a** dialog reappears with "Increase RV Value" or "Reconstruct STUDY" options, handled as described above. Successful pre-clustering updates the result label to "Pre-clustering completed successfully with 31 datasets and 335 dipoles." In the test example, pre-clustering processed 31 datasets and 335 dipoles, logging "Performing pre-clustering..." and validating dipole locations. Success updated the result label to "Pre-clustering completed successfully with 31 datasets and 335 components."

**Silhouette Analysis and Optimal Cluster Selection**

The plugin evaluates clustering viability by computing silhouette scores for 2 to 20 clusters using pre-clustered data and the selected algorithm (e.g., Kmeans, Optimal Kmeans, Affinity Propagation). For each cluster count, temporary clustering identifies assignments and outliers, calculating both mean and median silhouette scores (e.g., 0.31 mean and 0.29 median for 17 clusters). If clustering produces fewer clusters than requested or incomplete assignments, a dialog alerts the user, listing problematic cluster counts and offering two options: continue with the current algorithm or change the algorithm, resetting the app state. Results are visualized in three plots: a line plot of mean and median silhouette scores versus cluster numbers, with peaks marked for optimal counts (**Fig. S4b**); a grid of bar plots showing per-cluster mean scores with a mean line (**Fig. S4c**); and a boxplot of median silhouette scores across valid cluster counts, highlighting the best median score (**Fig. S4d**). The optimal cluster numbers based on mean and median scores (e.g., 17) are logged, the result label updates to "Silhouette analysis completed," and the "Number of Clusters" dropdown is enabled, allowing users to select the optimal number or another value. In the test example, silhouette analysis evaluated 2 to 20 clusters using the Kmeans algorithm, computing mean and median silhouette scores (e.g., 0.31 mean and 0.29 median for 17 clusters). The line plot (**Fig. S4b**) showed mean and median silhouette scores versus cluster numbers, with the optimal count (17) marked by a red star (mean) and red plus (median). Bar plots (**Fig. S4c**) displayed per-cluster mean scores with a red mean line, and the boxplot (**Fig. S4d**) showed median silhouette distributions across valid cluster counts, with the best median marked by a red plus. The plugin logged 17 as the optimal cluster count based on mean and median scores, updated the result label to "Silhouette analysis completed," and enabled the "Number of Clusters" dropdown for user selection.

**Final Clustering and Study File Saving**

Users select the number of clusters (e.g., 17, "Best Silhouette Score" based on mean or median) via the dropdown (**Fig. S3b**), which is stored in app.selectedNumClusters. For algorithms like Optimal Kmeans (without outlier handling) or Affinity Propagation, a dialog warns that the algorithm may not produce the exact number of clusters selected, offering options to continue with the current algorithm or change it, resetting the app state. Upon confirmation, the plugin performs final clustering using pop_clust with the chosen algorithm and threshold, logging the selection (e.g., "Using 17 clusters"). It validates assignments against the STUDY, ensuring consistency (e.g., 335 dipoles assigned). The clustered STUDY is saved as clustered.study, the status label updates to "Final clustering completed," and the result label shows "Clustered STUDY file saved successfully," enabling the "Load clustered EEG STUDY file" button. Mismatches halt processing with an error message. In the test example, selecting "Best Silhouette Score" (17 clusters) via the GUI (**Fig. S3b**) set app.selectedNumClusters to 17, logging "Using 17 clusters." Final clustering assigned 335 dipoles to 17 clusters, with the status label showing "Final clustering completed" and the result label confirming "Clustered STUDY file saved successfully." The plugin saved the STUDY as clustered.study and enabled the "Load clustered EEG STUDY file" button.

**Loading and Processing Clustered Study File**

When the user clicks the "Load clustered EEG STUDY file" button in the plugin's graphical interface (**Fig. S3b**), the system verifies the integrity of key variables, such as the number of datasets and dipoles (e.g., 31 datasets and 335 dipoles), ensuring they match and contain valid numeric data. If discrepancies or missing data are detected, the process halts, displaying an error in the interface. The system then locates and initializes the latest EEGLAB version without a graphical interface. It loads the clustered.study file from the user-specified directory (app.studyFilePath), halting with an error if the file is missing. Optionally, it retrieves MRI data (standard_mri.mat) from EEGLAB's dipfit plugin for dipole visualization. The system identifies the number of clusters (e.g., 17 clusters) in the loaded STUDY file, checking for an outlier cluster (assigned ID 0 if present, else cluster IDs start at 1). It validates data availability for scalp maps, spectra, event-related potentials (ERP), event-related spectral perturbations (ERSP), and inter-trial coherence (ITC) plots by confirming the presence of EEG data, independent component analysis (ICA) weights, and event information in each dataset. If all clusters lack components, the process stops; otherwise, it proceeds, noting any missing data for specific clusters. In the test example, for a dataset with 31 subjects and 335 dipoles, clicking the "Load clustered EEG STUDY file" button (**Fig. S3b**) successfully loaded the clustered.study file from the specified directory. The system confirmed that dataset and dipole counts matched and that subject and component indices were valid. EEGLAB initialized correctly, and the STUDY file revealed 17 clusters, with an outlier cluster assigned ID 0 if present; otherwise, cluster IDs began at 1. Data checks verified that most clusters had sufficient EEG, ICA, and event data for generating scalp maps, spectra, ERP, ERSP, and ITC plots.

The system creates a cluster table (clust_table) to map cluster assignments, with three columns: ClustersID (cluster number, 0 for outlier if present), SubjectID (subject index), and icID (component index). It extracts assignments from the STUDY file, processing the outlier cluster (if present) and real clusters by parsing names. Invalid or empty cluster data are skipped, and the table is sorted by ClustersID. The system validates subject indices against previously detected data (e.g., 31 subjects), halting if mismatches occur. It calculates the total number of unique subjects and converts group ranges (e.g., HS: [1:15], PS: [16:33] for two groups) from strings to numeric arrays, ensuring all values match detected subject indices. If ranges are invalid, the process stops. The finalized table is saved as clust_table.mat in the study directory, using a unique file path to avoid overwrites. In the test example, the system generated a cluster table with 335 rows and three columns. For 31 subjects, it validated subject indices and confirmed group ranges (e.g., one group: subjects 1–15; another: 16–33). If an outlier cluster existed, it was included with ID 0; otherwise, clusters were numbered from 1. The table was saved as clust_table.mat in the study directory.

The system extracts dipole data from the non-clustered STUDY file, validating that dataset and dipole counts match (e.g., 31 datasets, 335 dipoles) and halting if discrepancies or missing data are found. It iterates through datasets, skipping those without dipole models, and extracts coordinates (posx, posy, posz) and residual variance (rv). Rows with invalid coordinates are removed, and the process halts if no valid dipoles remain. The data form a table (dipXyz_table2) with 12 columns: Subjects (subject name), SubjectID, posx, posy, posz, rv, icID (component index), icaweights, icawinv, icaact (ICA data), chanlocs (channel locations), and srate (sampling rate). Subject names are populated from dataset metadata, and ICA data are validated for size consistency. If a dataset's channel count differs from 60, the system interpolates to a standard 60-channel layout, logging any failures. The table is validated against detected subject indices and total dipole counts, halting on mismatches, and saved as dipxyz_table2.mat using a unique file path. In the test example, for the 31 datasets, the system extracted 335 dipoles, including their spatial coordinates and residual variance. It created a table with 335 rows and 12 columns. Datasets with non-standard channel counts were interpolated to 60 channels, ensuring compatibility for plotting. The table was validated for consistency with prior data and saved as dipxyz_table2.mat.

The system combines cluster and dipole data into a table (dipxyz_clust_table) with 13 columns: ClustersID, SubjectID, icID, Subjects, posx, posy, posz, rv, icaweights, icawinv, icaact, chanlocs, and srate. It matches rows from clust_table with dipXyz_table2 based on subject and component indices, skipping unmatched entries. If the resulting table is empty or row counts mismatch, the process halts. Subject indices are updated using detected subject names and IDs (e.g., 31 subjects), stopping if any are not found. The table is saved as dipxyz_clust_table.mat with a unique file path. The system compares the table's row count (e.g., 335 rows) to the total components in the STUDY file (excluding the parent cluster), logging a warning if they differ. Upon success, the interface updates to display "Status: Clustered data processed successfully" and "Ready to generate plots and statistics," enabling the plotting and statistics button (**Fig. S3b**). On failure, an error message is shown. In the test example, the system combined cluster and dipole data into a table with 335 rows and 13 columns. If an outlier cluster was present, it was assigned ID 0; otherwise, clusters started at ID 1. Row counts matched the cluster table, and subject indices were updated. The table was saved as dipxyz_clust_table.mat. The interface updated to "Status: Clustered data processed successfully" and "Ready to generate plots and statistics," enabling the plotting button (**Fig. S3b**).

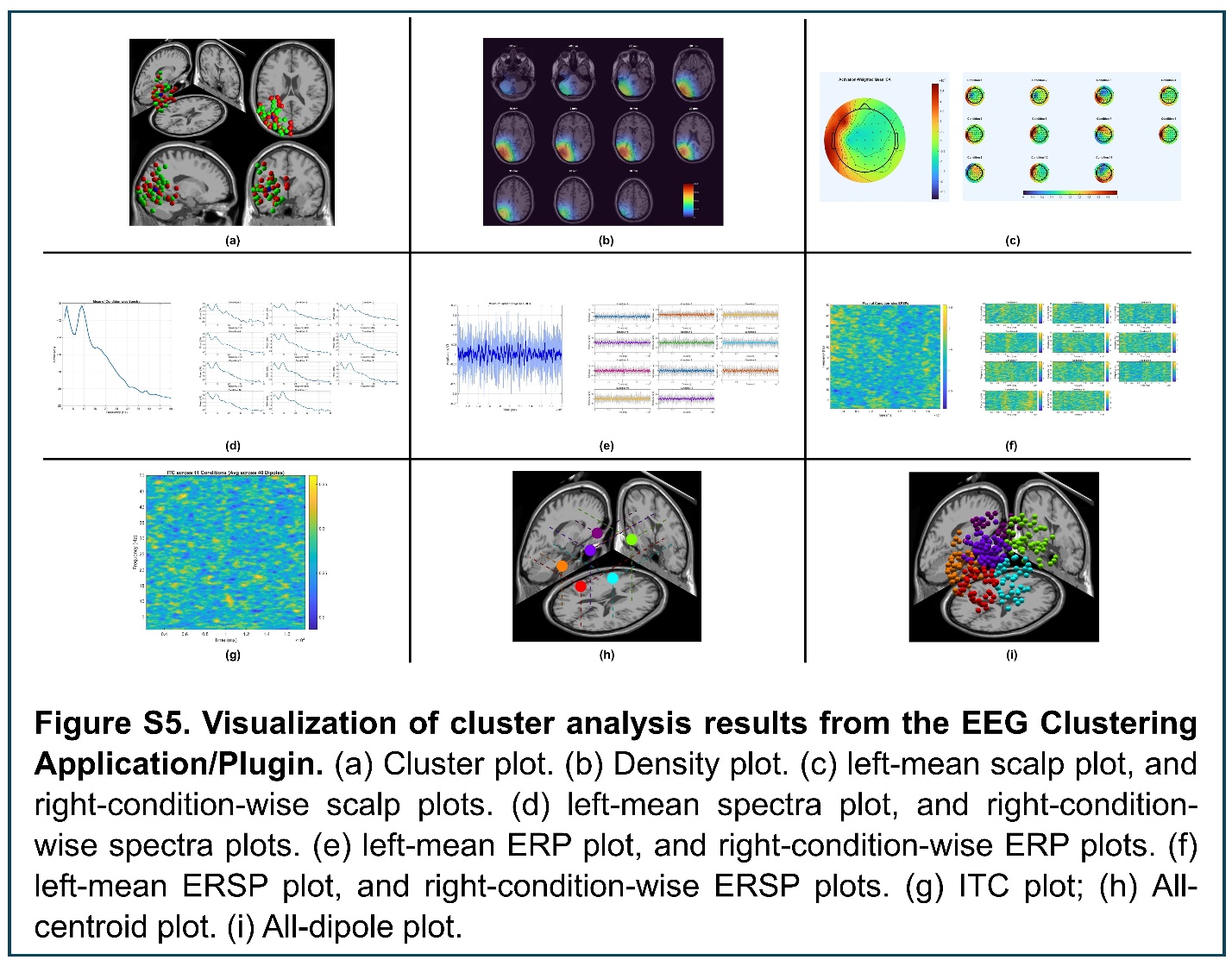

### 5. Visualization of cluster analysis results from the EEG Clustering Application/Plugin.

**Visualization and Statistical Analysis**

The EEG_Clustering plugin initiates visualization and statistical analysis by updating the user interface to display "Status: Generating cluster plots..." on the StatusLabel. Debug information, including group names (e.g., HS, PS for two groups), subject IDs, group prefixes, ranges (e.g., "1:13", "14:31"), and number of groups (1–3 supported), is logged for traceability and troubleshooting. The plugin dynamically identifies the latest EEGLAB version by searching the MATLAB path and selecting the most recent installation. If no valid EEGLAB is found, the process halts with an error displayed on the interface. The selected EEGLAB path is added, and EEGLAB is initialized in non-graphical mode to support automated processing of EEG data. A standard MRI file (standard_mri.mat) is loaded from the EEGLAB dipfit plugin directory to provide anatomical context for dipole visualizations. If unavailable, an alternative is located within the plugin's directory. The MRI data are validated for a valid mri structure, and compatibility with FieldTrip's ft_read_mri function is confirmed. Errors in loading halt the

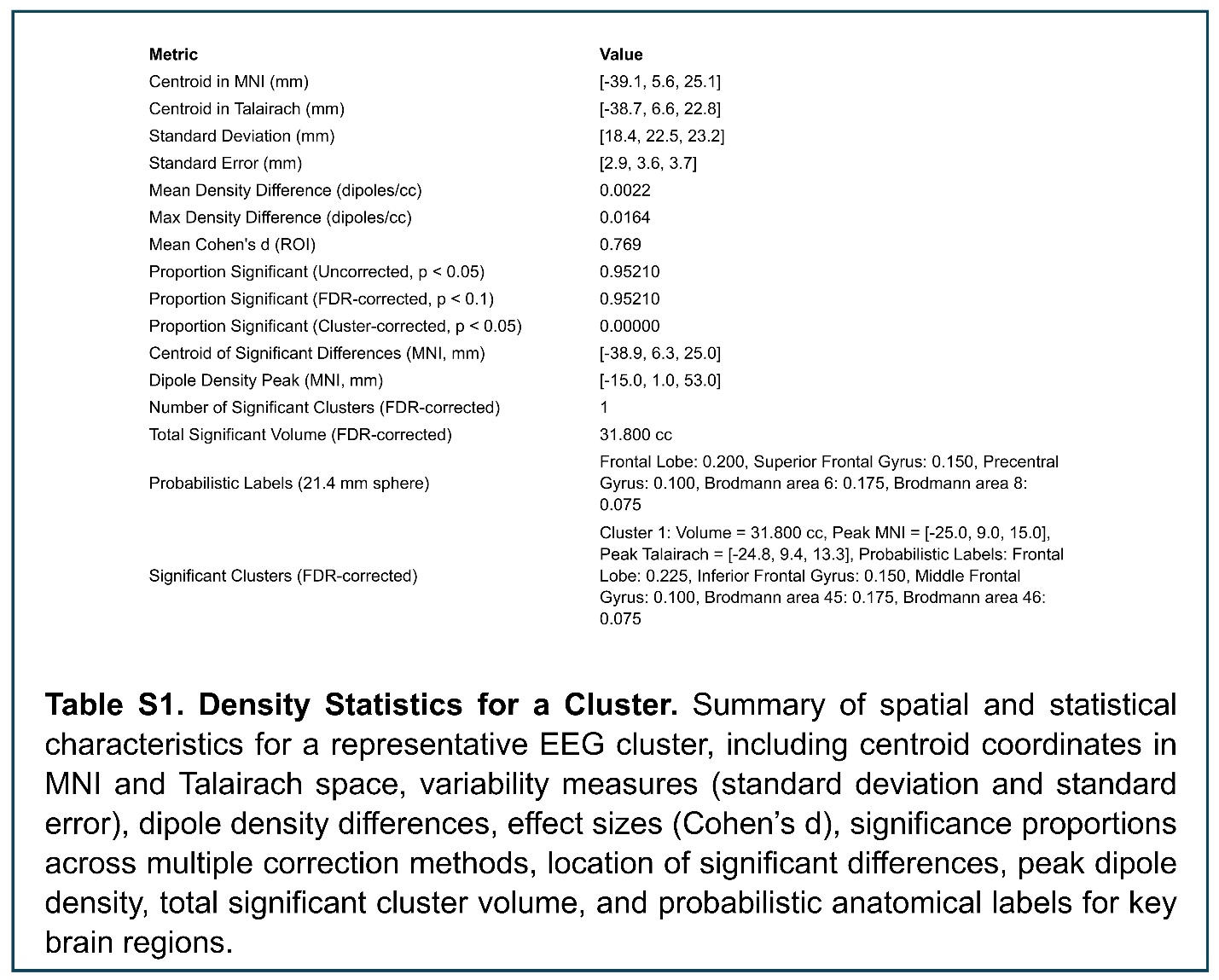

### 6. Density Statistics for a Cluster.

process, ensuring a reliable anatomical reference. In the test example, the plugin successfully initialized for a dataset with 31 subjects, 335 dipoles, and 17 clusters, divided into two groups (HS: 13 subjects, PS: 18 subjects). The StatusLabel updated to "Status: Generating cluster plots...", and debug logs recorded group names (HS, PS), subject IDs (1–31), group ranges ("1:13", "14:31"), and two groups. EEGLAB version was detected and initialized in non-graphical mode, and the standard_mri.mat file was loaded from the EEGLAB dipfit directory and validated. Cluster IDs are extracted from dipxyz_clust_table, identifying unique clusters (e.g., 17, including an outlier cluster with ID 0 if present). Processing starts at ID 0 with outliers or ID 1 otherwise. If no clusters are detected, a warning halts the process. Subject IDs (e.g., 31 unique subjects) are validated, and group ranges (e.g., "1:13" for HS, "14:31" for PS) are parsed into numeric intervals. Assigned subjects are verified against unique subjects to ensure consistency, with mismatches triggering errors. If an outlier cluster (ID 0) exists, its data are extracted from dipxyz_clust_table and saved as outlier_cluster.mat in the study directory. A dipole plot is generated using MNI coordinates, with dipoles colored by group (e.g., green for HS, red for PS, blue for the centroid) overlaid on MRI data. The plot is saved with a timestamped filename. Errors, such as an empty cluster, update the interface with a failure message and halt processing. In the test example, seventeen unique cluster IDs (0–16, including an outlier cluster) were extracted from dipxyz_clust_table. Processing started at ID 0, with 31 unique subjects assigned to groups (13 to HS, 18 to PS) using ranges "1:13" and "14:31". An outlier cluster (ID 0) containing approximately 10 dipoles was processed and saved as outlier_cluster.mat. A dipole plot was generated, displaying dipoles in green (HS), red (PS), and blue (centroid) overlaid on MRI data and saved. If multiple groups are detected (e.g., HS, PS), the plugin prompts the user to select an analysis type via a dialog: group-combined analysis, group-wise analysis, or both. Selecting group-combined analysis (default) or both triggers analysis across all groups combined. For real clusters (IDs ≥ 1), a group-combined directory is created with subfolders for each cluster (e.g., Cluster_1 to Cluster_17). Data structures store dipole positions, ICA activations, spectra, ERPs, ERSPs, and ITCs for 2–20 clusters. Dipole positions and sampling rates are extracted, centroids are computed in MNI and Talairach coordinates, and colors are assigned (e.g., HSV colormap for clusters). For each cluster, the plugin generates the following visualizations and statistics: (1) Cluster Plot (**Fig. S5a**): Displays the spatial distribution of dipoles in MNI coordinates, with group-specific coloring (e.g., green for HS, red for PS) to highlight differences in dipole locations across groups. A blue centroid marks the cluster's central tendency, overlaid on MRI anatomy to contextualize neural source locations. (2) Dipole Density Plot (**Fig. S5b**): Illustrates the three-dimensional spread of dipoles across each cluster, highlighting the concentration and dispersion of neural sources, which supports the evaluation of cluster integrity and spatial organization. (3) Scalp Topoplots (**Fig. S5c**): Shows mean and condition-wise EEG activity projected onto a 2D scalp map, revealing spatial patterns of neural activity (e.g., voltage distributions) across electrodes for each condition, facilitating comparison of topographic differences. (4) Spectra Plots (**Fig. S5d**): Depicts mean and condition-wise power spectra (1–50 Hz), illustrating frequency-specific neural activity (e.g., alpha, beta bands) to identify oscillatory differences across conditions or groups. (5) ERP Plots (**Fig. S5e**): Presents mean and condition-wise event-related potentials with 50-ms smoothing, showing time-locked EEG responses to stimuli, enabling analysis of temporal dynamics and amplitude differences. (6) ERSP Plots (**Fig. S5f**): Displays mean and condition-wise event-related spectral perturbations (1–50 Hz), highlighting time-frequency changes in power to reveal dynamic oscillatory responses to experimental conditions. (7) ITC Plot (**Fig. S5g**): Shows inter-trial coherence averaged across conditions and dipoles, quantifying phase consistency of neural responses across trials, useful for assessing synchronization in event-related activity. (8) All-Centroid Plot (**Fig. S5h**): Visualizes centroids of all clusters in MNI coordinates, colored by cluster (HSV colormap), to compare spatial relationships and anatomical localization across clusters. (9) All-Dipole Plot (**Fig. S5i**): Displays all dipoles across clusters, colored by group, to provide a comprehensive view of dipole distributions and group differences in neural source locations.

Density Statistics (**Table S1**): Computed using 5000 permutations to generate a non-parametric distribution of dipole locations, yielding centroids (mean dipole positions), standard deviations (spatial variability within clusters), and FDR-corrected p-values (p < 0.1) to assess statistical significance of spatial differences between clusters. Low p-values indicate distinct neural sources, while standard deviations quantify cluster compactness. Probabilistic Talairach labels assign anatomical brain regions to centroids (e.g., frontal lobe, temporal cortex), providing functional context for neural activity. Results are saved, and all data are stored. The visualization format applies consistently across all clusters. In the test example, with two groups detected (HS, PS), the plugin prompted the user to select an analysis type, and group-combined analysis was chosen. Sixteen real clusters (IDs 1–16) with 325 dipoles were analyzed, with data structures storing dipole positions, ICA activations, spectra, ERPs, ERSPs, and ITCs. Visualizations for each cluster (e.g., Cluster 1, **Fig. S5A-I**) included cluster plots (**Fig. S5a**), dipole density plots (**Fig. S5b**), mean and condition-wise scalp topoplots (**Fig. S5c**), spectra (**Fig. S5d**), ERPs (**Fig. S5e**), ERSPs (**Fig. S5f**), and ITCs (**Fig. S5g**) generated and saved. Density statistics (**Table S1**) reported centroids, standard deviations, p-values (FDR-corrected, p < 0.1), and probabilistic Talairach labels, and saved. All-centroid (**Fig. S5h**) and all-dipole (**Fig. S5i**) plots visualized all 16 clusters and their associated dipoles, with data and plots saved.

**Interpretation of Density Statistics (Table S1):**

- **Centroid in MNI and Talairach:** The centroid coordinates [-39.1, 5.6, 25.1] in MNI space and [-38.7, 6.6, 22.8] in Talairach space represent the average location of dipoles in the cluster, indicating a primary neural source in the left frontal region. These coordinates allow mapping to brain anatomy for functional interpretation.
- **Standard Deviation:** Values [18.4, 22.5, 23.2] mm across the x, y, and z axes reflect the spatial spread of dipoles, suggesting moderate variability in dipole locations, with the largest dispersion along the z-axis (depth).
- **Standard Error:** [2.9, 3.6, 3.7] mm indicates the precision of the centroid estimates, showing that the mean dipole location is reliably estimated with low uncertainty.
- **Mean and Max Density Difference:** The mean density difference of 0.0022 dipoles/cc and max difference of 0.0164 dipoles/cc quantify the average and peak variation in dipole concentration within the cluster, highlighting regions of higher neural activity density.
- **Mean Cohen's d (ROI):** A value of 0.769 indicates a moderate to large effect size of dipole density differences within the region of interest (ROI), suggesting a meaningful distinction in neural activity between groups or conditions.
- **Proportion Significant (Uncorrected, p < 0.05):** A value of 0.95210 shows that 95.21% of the cluster's dipole locations are statistically significant without multiple comparison correction, indicating strong initial evidence of clustering.
- **Proportion Significant (FDR-corrected, p < 0.1):** Also 0.95210, this confirms that after controlling for false discovery rate, 95.21% of locations remain significant, reinforcing the robustness of the clustering at a 10% FDR threshold.
- **Proportion Significant (Cluster-corrected, p < 0.05):** A value of 0.00000 indicates that no locations survive a stricter cluster-level correction, suggesting caution in interpreting broad spatial significance without further validation.
- **Centroid of Significant Differences:** [-38.9, 6.3, 25.0] in MNI space pinpoints the average location of statistically significant dipole differences, closely aligning with the overall centroid and reinforcing the frontal focus.
- **Dipole Density Peak:** [-15.0, 1.0, 53.0] marks the highest concentration of dipoles, located higher and more medial than the centroid, possibly indicating a secondary active region.
- **Number of Significant Clusters and Total Significant Volume:** One significant cluster with a volume of 31.800 cc (FDR-corrected) defines the spatial extent of reliable dipole activity, providing a measure of the cluster's anatomical footprint.

**Probabilistic Labels:** Within a 21.4 mm sphere, probabilities (e.g., Frontal Lobe: 0.200, Superior Frontal Gyrus: 0.150) estimate the likelihood of the centroid residing in these regions, with Brodmann areas 6 (0.175) and 8 (0.075) suggesting involvement in motor planning and eye movement control.

- **Significant Clusters:** The cluster's 31.800 cc volume, with a peak at [-25.0, 9.0, 15.0] (MNI) and labels like Inferior Frontal Gyrus (0.150) and Brodmann area 45 (0.175), indicates a significant frontal activation zone, potentially linked to language or cognitive processing.

When multiple groups are detected (e.g., HS, PS), the user is prompted to choose group-combined analysis, group-wise analysis, or both. Selecting group-wise analysis or both initiates group-specific analysis. For a single group, this step runs automatically. For each group (e.g., HS, PS), a group-specific directory is created with cluster subfolders. Data are filtered by group-specific subject IDs (e.g., 13 for HS, 18 for PS), and analyses mirror the combined analysis described above. All the visualizations and statistics follow the same pattern. In the test example, for the two groups (HS, 13 subjects, ~170 dipoles; PS, 18 subjects, ~165 dipoles), the user selected group-wise analysis or both via the analysis type dialog. For HS (13 subjects, ~170 dipoles) and PS (18 subjects, ~165 dipoles), group-specific directories were created, and data were filtered by subject IDs. Each group's 16 clusters were analyzed, producing visualizations and statistics mirroring the combined analysis, with all plots and data saved in group specific directories.

Upon completion, the StatusLabel updates to "Status: Clustered data processed successfully," and the ResultLabel confirms readiness. A green progress bar indicating 100% completion is displayed for 5 seconds. A summary log details total clusters, real clusters, outlier presence, and groups analyzed. In the test example, the StatusLabel updated to "Status: Clustered data processed successfully," and the ResultLabel confirmed readiness. A green progress bar indicating 100% completion was displayed for 5 seconds. The summary log reported 17 clusters (16 real, 1 outlier) and 2 groups (HS, PS), with processing completing without errors.

### 7. Pooled and tier-specific distributions of fitted-dipole depth

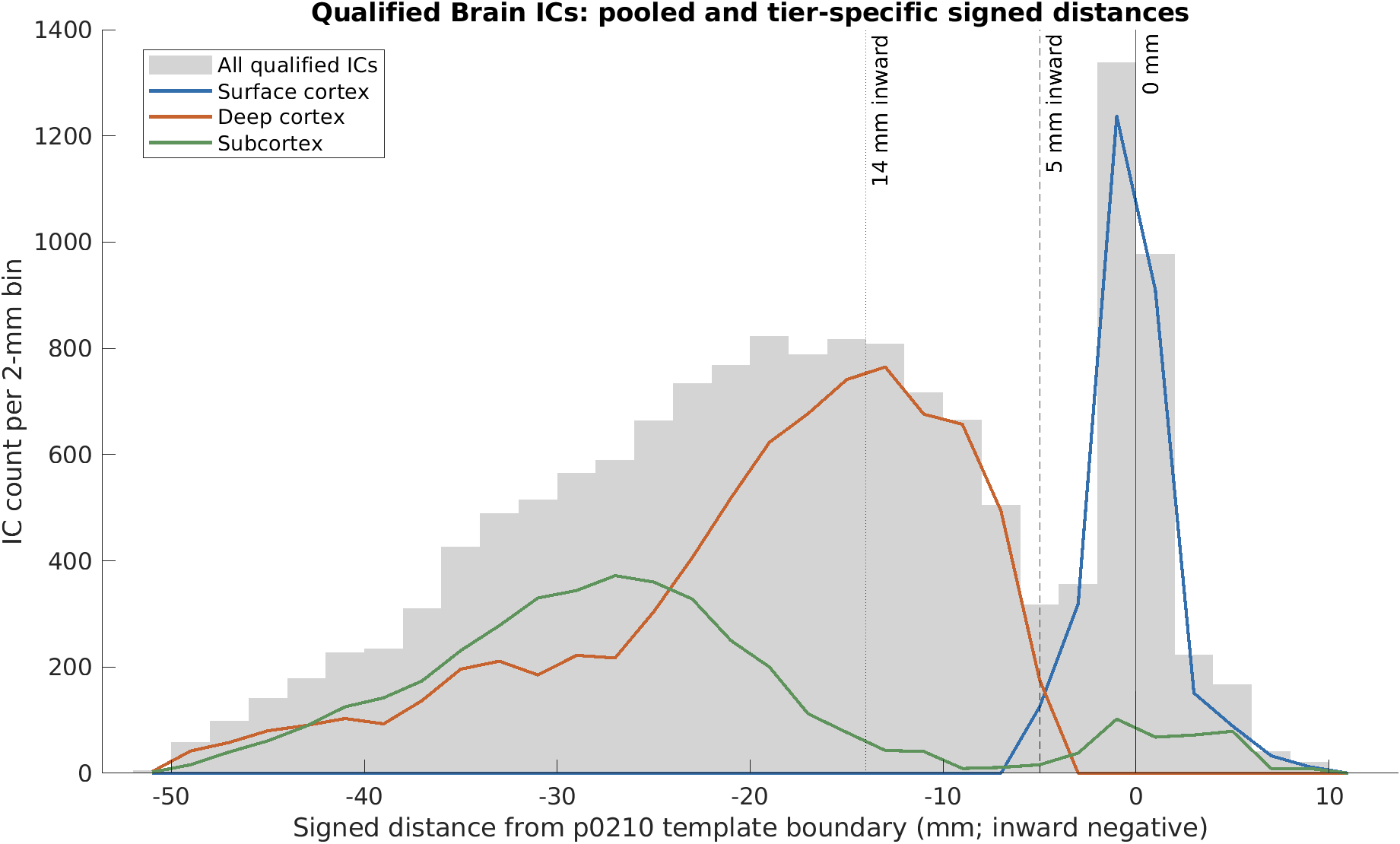

**Figure S7. Pooled and tier-specific signed-distance distributions of qualified Brain ICs.** Gray bars show the pooled distribution of all 14,578 qualified Brain ICs, and colored lines show the distributions for the three operational spatial tiers: Surface cortex (n = 2,873), Deep cortex (n = 7,656), and Subcortex (n = 4,049). Signed distances were measured from the template boundary derived from the EEGLAB DIPFIT head model, with negative values indicating inward locations and positive values indicating outward locations. All histograms use 2-mm bins. The pooled distribution exhibits two prominent peaks near −1 mm and −19 mm, separated by a pronounced minimum near −5 mm. Vertical lines indicate the template boundary (0 mm), the 5-mm inward classification threshold, and the 14-mm inward reference distance. The operational tiers were defined using distance, anatomical labels, and an inferior-coordinate criterion, as described in the main text.
